# Impaired regulatory T-cell–mediated immune tolerance promotes neurodegeneration in glaucoma

**DOI:** 10.64898/2026.09.17.26363354

**Authors:** Huilan Zeng, Merri-Grace Jones, Zeb R. Zacharias, Erin A. Boese, Wallace L.M. Alward, Young H. Kwon, Jon C.D. Houtman, Edward Linton, Randy H. Kardon, Oliver W. Gramlich, Markus H. Kuehn

**Affiliations:** Department of Ophthalmology and Visual Sciences, The University of Iowa; Department of Ophthalmology, The First Affiliated Hospital, Hengyang Medical School, University of South China; Veterans Affairs Center for the Prevention and Treatment of Visual Loss, Iowa City VA Health Care System; Human Immunology Core, The University of Iowa; Department of Microbiology and Immunology, The University of Iowa; Holden Comprehensive Cancer Center, University of Iowa; Department of Ophthalmology and Visual Sciences, University of Alabama at Birmingham

## Abstract

Neurodegenerative diseases are increasingly recognized to involve detrimental interactions between the immune and nervous systems, yet whether failure of peripheral immune tolerance actively drives neuronal loss frequently remains unclear. Here, using primary open-angle glaucoma as a model of chronic neurodegeneration, we demonstrate that dysregulated adaptive immunity is sufficient to promote retinal ganglion cell degeneration. Peripheral blood mononuclear cells from glaucoma patients, but not healthy donors, induced retinal ganglion cell loss following transfer into humanized immunodeficient mice without changes to the intraocular pressure, demonstrating a causal role for immune responses in the patient derived material. Comprehensive immune profiling further revealed selective changes in the regulatory T-cell compartment, indicating reduced activation, an impaired suppressive phenotype, and altered differentiation and trafficking states despite preserved overall regulatory T-cell abundance. We further demonstrate that transient expansion of regulatory T cells preserves visual function, reduced optic nerve axonal degeneration, and limited retinal ganglion cell loss in an experimental glaucoma model. Together, these findings identify failure of regulatory T-cell–mediated immune tolerance as a mechanism that permits neurodegeneration in glaucoma and demonstrate that restoring immune regulation can ameliorate neuronal injury. Our data indicate that immune tolerance is a modifiable determinant of chronic neurodegeneration and suggest that immunoregulatory therapies may complement conventional pressure-lowering treatments to preserve vision in glaucoma.

## Introduction

Age-related neurodegenerative diseases are a leading cause of disability worldwide and are characterized by the progressive loss of neuronal structure and function. These disorders have traditionally been viewed as neuron-centered diseases, but growing evidence indicates that immune mechanisms are integral to disease pathogenesis as well as progression and that extensive interactions between the nervous and immune systems shape neuronal survival and repair (1, 2). Studies in Alzheimer’s disease, Parkinson’s disease, and other age-related neurodegenerative disorders have identified alterations in adaptive immune responses, including changes in T-cell activation, clonal expansion, and extravasation into the central nervous system (3–6). In the healthy central nervous system mechanisms of peripheral immune tolerance restrain autoreactive lymphocytes and protect neural tissues from immune-mediated injury. However, aging is associated with immunosenescence and chronic low-grade inflammation and these processes may disrupt immune homeostasis and compromise regulatory pathways that normally maintain self-tolerance (7, 8). Elucidating how age-associated dysfunction of adaptive immunity contributes to neurodegeneration is therefore of considerable importance. Here, we investigate the mechanisms by which altered immune regulation promotes chronic neuronal loss and evaluate the therapeutic potential of restoring immune tolerance as a strategy to preserve neuronal integrity and function (9).

The glaucomas are a group of optic nerve diseases that together are the second leading cause of irreversible blindness worldwide (10). Elevated intraocular pressure (IOP) is a significant risk factor for the development of glaucoma (11), but the susceptibility to IOP elevation varies among individuals, and a number of additional factors undoubtedly affect vision loss in glaucoma (12–14). It is becoming increasingly evident that retinal neuroinflammation is one of the factors that contribute to retinal ganglion cell (RGC) degeneration, leading to loss of neuronal signal transduction from the eye to the brain which is the ultimate cause of vision loss (15). The neuroinflammatory response is mainly characterized by the activation of macroglial and microglia cells within the neuroretina. Increased levels of the cytokines TNF and IL-1β, activation of the complement cascade, and the formation of inflammasomes are all well-established events in the glaucomatous retina (16–19). However, recent results from multiple laboratories indicate that adaptive immunity also contributes to glaucomatous RGC damage and these studies collectively suggest that T-cells may play a far more significant role in the pathogenesis of the disease than previously assumed (20–22).

Our published studies have demonstrated that lymphocytes are at least partially responsible for RGC loss in animal models of glaucoma. Experimental induction of ocular hypertension in Rag1-/- mice, which lack mature T and B lymphocytes, results in approximately 50% less RGC attrition compared to immunocompetent controls. Moreover, unilateral induction of elevated IOP causes mild RGC loss in the contralateral eye in normal mice but not in Rag1^-/-^, indicating that RGC damage in contralateral eyes is largely immune mediated(23). In a second study, we carried out adoptive transfers of immune cell populations from mice with glaucoma into naïve recipients. These transfers did not lead to IOP elevation in recipient mice, but mice that received either total splenocytes or T cells did experience progressive loss of RGC during the following 16 weeks, demonstrating the presence of autoreactive lymphocytes in these glaucoma models (24).

In addition to elevated IOP, advanced age is also a primary risk factor for the development of glaucoma (25–27). Advanced age is also associated with impaired regulation of inflammation and age related changes include chronic upregulation of inflammatory cytokines, and altered immune populations (28). T-regulatory cells (Tregs) are essential for the induction and maintenance of peripheral immune tolerance through the suppression of self-reactive immune cells and excessive inflammation (29–31), but the function of Tregs frequently becomes compromised with age (32–35). Several studies have reported differences in the distribution of T cell subtypes between controls and glaucoma patients, including increased frequencies of CD3+ T cells and Th1 T cells and increased expression of the soluble interleukin-2 (IL-2) receptor, a cytokine required for T cell proliferation (18, 36, 37). Peripheral immune profiling demonstrates altered Treg frequencies in glaucoma patients, although the direction of change is variable across studies (36–38). This could be attributed to differences in Treg phenotypic definitions and gating approaches, or it might reflect variances in disease stage or immune activation status. Ultimately, a more in depth characterization of Tregs in glaucoma is warranted.

In the present study, we aimed to determine whether a T cell compartment imbalance can also be observed in POAG patients and to functionally demonstrate whether this altered distribution contributes to RGC loss. Toward this end, we transferred peripheral blood mononuclear cells (PBMC) from POAG patients and eye disease-free controls into NOD/scid gamma (NOD.Cg-*Prkdc ^scid^ Il2rg^tm1Wjl^*/SzJ, NSG) mice. NSG mice are commonly used to model human autoimmune and chronic inflammatory diseases (39) and in our hands NSG mice that received PBMC from patients experienced significantly more RGC loss when compared to those having received PBMCs from healthy donors. We further conducted an in depth immunophenotyping of PBMC obtained from patients with POAG and healthy controls. To our knowledge, this study comprises the largest POAG cohort analyzed to date and represents the most comprehensive immunophenotyping of glaucoma reported thus far.

Our data indicate that the overall T cell profile in POAG represents an “early effector phenotype” favoring a pro-inflammatory milieu. This pro-inflammatory signature appears partially related to a loss of Treg function, which limits their capacity to suppress other T cell populations. Finally, we demonstrate that expansion of Tregs in the context of elevated IOP lessens visual loss and protects against optic nerve axonal damage and RGC loss in an experimental glaucoma model. Taken together, this work demonstrates that an imbalanced T cell distribution, driven by Treg dysfunction, occurs in POAG patients and contributes directly to RGC degeneration.

## Results

### Adoptive transfer of POAG-derived PBMCs exacerbates RGC loss following IOP elevation

We hypothesized that immune-mediated mechanisms contributing to glaucomatous neurodegeneration are most pronounced during phases of active disease progression. Optic disk hemorrhages often signal ongoing optic nerve damage and are associated with disease progression, even in patients who appear stable (40, 41). Therefore, peripheral blood mononuclear cells (PBMCs) were collected from POAG patients (n=7) with apparent optic disk hemorrhage at examination to maximize the likelihood of detecting relevant immune signatures. PBMC were also obtained from healthy controls (n=10) without glaucoma or signs of optic disk hemorrhage and 3.5 - 5 x 10^6^ cells were immediately injected intraperitoneally into NSG mice (Figure 1 A).

**Figure 1:**
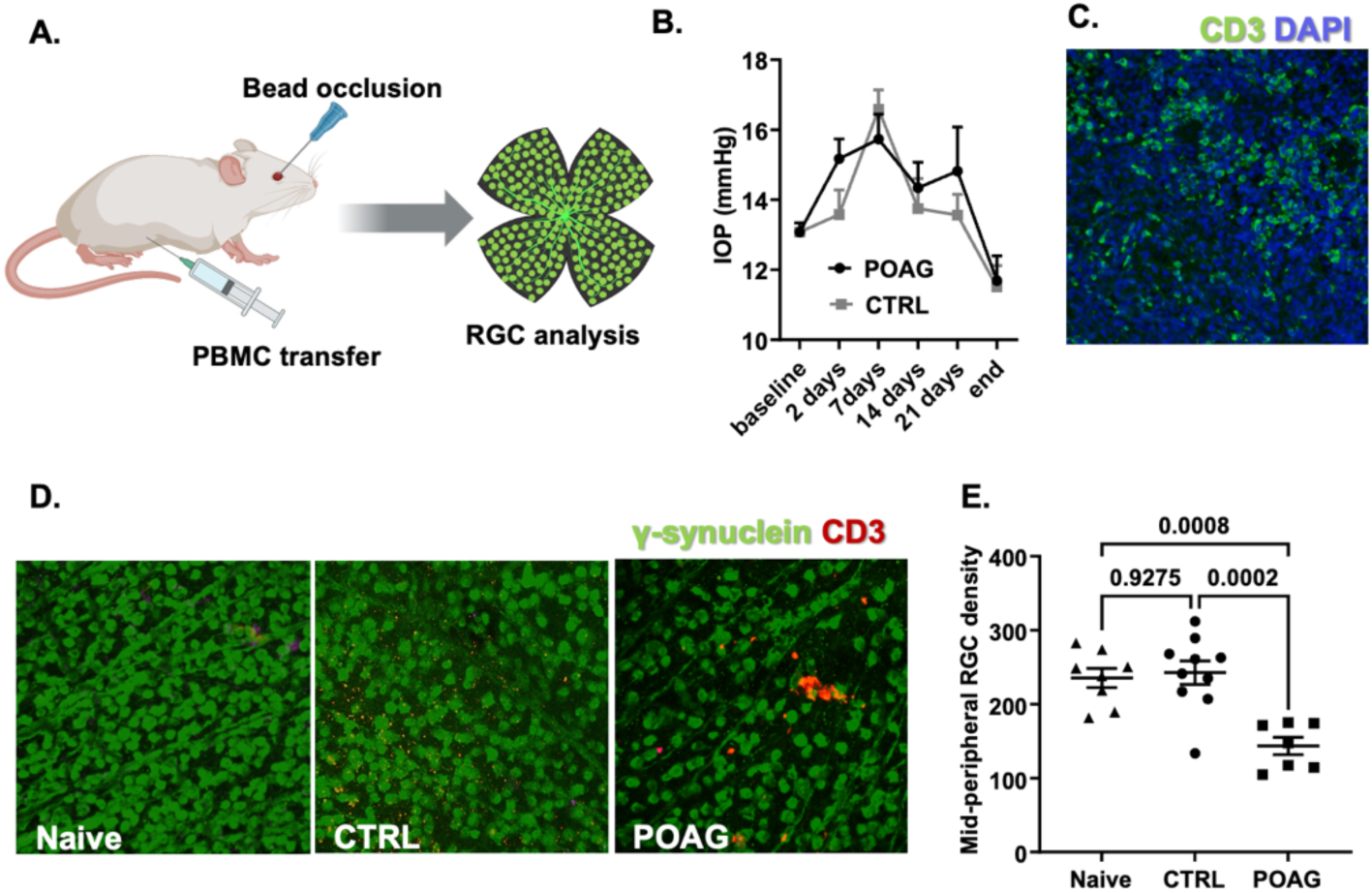
Adoptive Transfer of POAG-Derived PBMCs Exacerbates RGC Loss Following IOP Elevation. (A) Schematic of the experimental design. PBMCs from POAG patients (n=7) or healthy controls (n=10) were transferred into NSG mice, followed 2 weeks later by microbead-induced IOP elevation. (B) IOP after microbead injection. POAG and control PBMC-recipient mice developed comparable mild IOP elevation within 5–7 days. (C) Human CD3+ T-cell (green) engraftment in the spleen of an NSG mouse 54 days after PBMC transfer. (D) Representative retinal whole mounts from naïve (no PBMC, no microbeads), CTRL (healthy donor PBMC), and POAG (POAG donor PBMC) mice stained for γ-synuclein and human CD3. (E) RGC density quantification. Mice receiving healthy PBMCs showed no RGC loss versus naïve NSG mice (p=0.9275). In contrast, mice receiving PBMCs from POAG patients with disc hemorrhage exhibited reduced RGC density compared with both naïve NSG mice (p=0.0008) and healthy donor recipients (p=0.0002).

After an engraftment period of two weeks, a mild IOP challenge was induced bilaterally using the microbead occlusion model (42). The amount of microbeads injected was carefully chosen to induce only minor and temporary IOP elevation that will not result in RGC loss. Using this approach, both POAG PBMC-recipient and healthy control PBMC-recipient mice developed comparable IOP elevations within 5–7 days after injection, with mean peak IOP values of 16.9 ± 1.9 mmHg and 16.7 ± 1.8 mmHg, respectively (Figure 1B). These IOP values were only slightly higher than baseline and returned to baseline levels within one week thereafter. Importantly, there was no significant difference in IOP elevation between POAG and control PBMC-recipient mice

Animals were sacrificed 54 days after PBMC transfer and systemic engraftment of CD3 positive cells was confirmed in the spleens of all NSG mice, regardless of donor status (Figure 1 C). In the absence of endogenous T cells in these mice, all CD3+ cells must be derived from the transplanted human PBMC. We then determined the density of γ-synuclein+ RGC in retinal whole-mount preparations in NSG mice that received neither PBMC transfer nor microbead injections (Naïve), PBMC from healthy controls and microbead injections (CTRL), and those that transfer of PBMC from POAG donors with disk hemorrhage and microbead injections (Figure 1 D).

Quantitation of mid-peripheral RGC densities revealed that mice that received PBMC from healthy controls (CTRL) retained as many RGC as naive NSG animals (243±50 RGC/field vs. 235±37 RGC/field, p=0.93; Figure 1 E). In contrast, NSG mice that received transfers of PBMC from POAG donors with disk hemorrhage experienced a significant reduction in RGC density (144±31 RGC/field, 40.8% loss) when compared to CTRL mice (p=0.0002) or Naïve animals (p=0.0008, Figure 1 E). Examination of these retinas also revealed a small number of T cells in the midperipheral region of the retinas (Figure 1D). However, no apparent differences in the number of T cells or their location within the retina was observed between the cohorts.

### Immune cell mediated RGC loss is correlated with glaucoma patient ophthalmic metrics

These findings align with those obtained in previous studies using mouse-to-mouse transfers (23, 24) and suggest that adaptive immunity mediated damage mechanisms could also contribute to vision loss in some patients. Furthermore, the majority of POAG patients do not display disk hemorrhages at the time of examination and furthermore very strict exclusion criteria were applied during patient recruitment during this first phase of the study which could potentially skew the observed responses. In order to assess whether immune mediated RGC loss is observed in a wider range POAG patients, we repeated the transfer experiments with PBMC obtained from POAG patients without signs of disk hemorrhage. As before, PBMC were obtained from POAG patients (n=17) or healthy controls (n=11) and transferred into NSG mice by intraperitoneal injection. Two weeks after transfer, IOP elevation was induced bilaterally as described above. RGC density analysis was conducted 50 days after PBMC transfer (36 days after IOP challenge). To gain additional information regarding the regional distribution of RGC loss, RGC loss was evaluated in the peripheral, mid-peripheral and central regions of the mouse retina. The resultant data demonstrate that NSG mice that received PBMC from POAG donors displayed significantly fewer RGC in the central area of the retina than those receiving PBMC transfer from controls (255±65 RGC/field vs. 341±131 RGC/field, -25.2%, p=0.034, Figure 2 A). A similar effect was not observed in the peripheral region of the retinas (-10.9%, p=0.44), while mid-peripheral regions displayed an intermediate damage phenotype (-24.1%, p=0.145).

**Figure 2.**
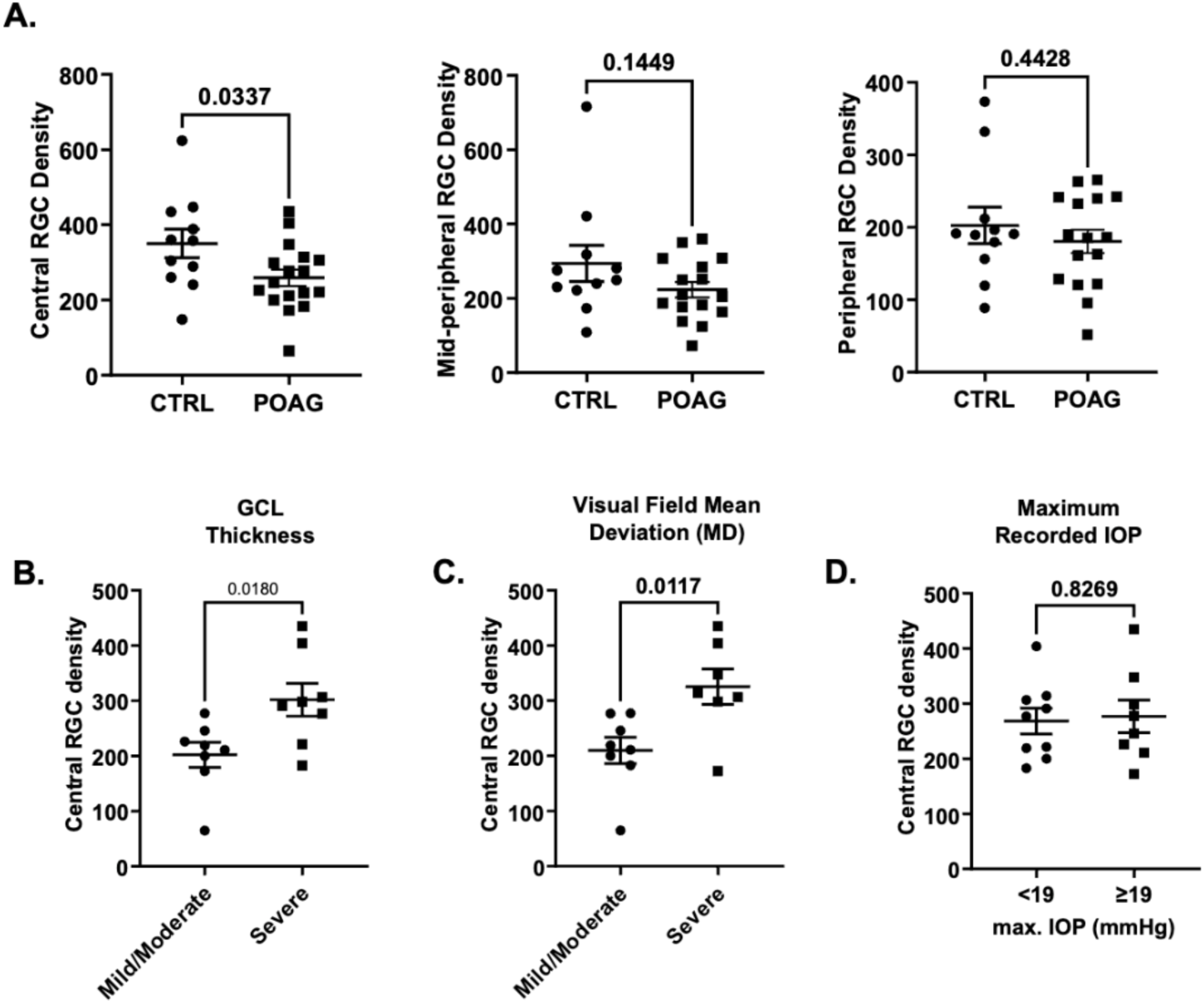
Immune Cell–Mediated RGC Loss in Recipient Mice Correlates with Glaucoma Patient Ophthalmic Metrics. (A) RGC density in the central (p = 0.0337), mid-peripheral (p = 0.1149), and peripheral retina (p = 0.4428) of mice receiving PBMCs from POAG patients. Central RGC density in POAG PBMC-recipient mice after stratifying POAG donors according to clinical metrics. (B) Recipients of PBMCs from donors with severe GCL thinning exhibited higher central RGC density than recipients of PBMCs from donors with mild to moderate GCL thinning (p = 0.0180). (C) A similar pattern was observed when donors were stratified by visual field mean deviation, with PBMCs from donors with more severe visual field loss inducing less RGC loss in recipient mice (p = 0.0117). (D) In contrast, stratification by maximum recorded IOP revealed no difference in central RGC density (p = 0.8269). All statistical analyses were performed using an unpaired Student’s t-test.

To gain additional insight into patterns of RGC loss relevant to clinical parameters of POAG, we compared the severity of central RGC loss observed in NSG mice with clinical measures from corresponding human donors, including maximum recorded IOP, visual field mean deviation (MD), and macular ganglion cell layer (GCL) thickness. GCL thickness reflects the loss of RGCs and their axons and therefore serves as a structural indicator of glaucomatous damage, whereas MD provides a functional assessment of visual sensitivity relative to healthy, age-matched individuals and declines with disease progression. These clinical data were available only for POAG donors, and our aim was to determine whether RGC loss in NSG mice corresponded to clinical indicators of mild-to-moderate versus advanced glaucoma in the donor cohort. For this analysis, POAG donors were stratified into upper and lower halves for each parameter: GCL thickness above or below 60 µm in the worse eye, MD greater or less than -8 dB in the worse eye, and maximum recorded IOP above or below 19 mmHg in the worse eye. It should be noted, however, that all POAG patients had received IOP-lowering therapy, and their untreated IOP levels was therefore unknown (Table 1). Further, the POAG cases with advanced disease exhibited the lowest IOP on average, presumably due to more aggressive treatment in this cohort.

**Table 1.** Demographic and clinical characteristics of study participants. . Intraocular pressure (IOP), retinal nerve fiber layer (RNFL) thickness, and ganglion cell layer (GCL) thickness were measured on the day of sample collection. Reported values represent the means of the worse eye for each participant within each group. Visual field loss was assessed only in POAG patients and is reported in decibels as the mean of the worse eye for each participant. All POAG patients were receiving IOP-lowering treatment at the time of assessment. F: Female, M: Male, C: Caucasian, H: Hispanic, Af: African, As: Asian, O: Other/Unknown

|  | Age | Sex | Ethnicity | IOP | RNFL Thickness | GCL Thickness | Mean Deviation |
| --- | --- | --- | --- | --- | --- | --- | --- |
| **Controls**<br>**(n=38)** | 72.2 ± 8.3 years | F: 22.5%<br>M: 77.5% | C: 78.9%<br>H: 7.89%<br>Af: 2.63%<br>As: 0%<br>O: 10.52% | 15.8 ± 3.0 mmHg | 83.0 ± 13.0 µm | 69.5 ± 14.8 µm | N/A |
| **Mild/Moderate POAG**<br>**(n=19)** | 70.6 ± 9.2 years | F: 30.4%<br>M: 69.6% | C: 95.6%<br>H: 0%<br>Af: 0%<br>As: 0%<br>O: 4.34% | 15.7 ± 3.3 mmHg | 66.5 ± 10.6 µm | 60.3 ± 11.2 µm | -4.1 ± 3.2 dB |
| **Severe POAG**<br>**(n=27)** | 75.5 ± 8.1 years | F: 25.8%<br>M: 74.2% | C: 63.63%<br>H: 3.0%<br>Af: 6.06%<br>As: 3.0%<br>O: 24.3% | 13.3 ± 4.2 mmHg | 55.9 ± 16.0 µm | 45.0 ± 22.9 µm | -10.0 ± 6.3 dB |
| **P value** | ANOVA: 0.11<br>Brown-Forsythe: 0.90 | Chi Square: 0.78 | Chi Square: 0.18 | ANOVA: 0.01<br>Brown-Forsythe: 0.03 | ANOVA: 0.011<br>Brown-Forsythe: 0.76 | ANOVA: 0.03<br>Brown-Forsythe: 0.29 | T-Test: 0.009 |

Our analysis revealed that transfer of PBMCs from donors with severe GCL thinning resulted in better RGC survival in recipient mice than transfer of PBMCs from donors with mild to moderate disease (Figure 2B, p=0.018). A similar pattern was observed when stratifying patients by visual field status: PBMCs from POAG donors with more vision loss induced significantly less RGC loss in NSG mice than PBMCs from donors with less advanced disease (p=0.012) (Figure 2 C). In contrast, a difference between higher or lower max IOP values was not observed (Figure 2 C, p=0.82). Together, these findings indicate that immune-mediated mechanisms contributing to RGC loss in POAG are more prominent during moderate stages of disease than in advanced glaucoma.

### Altered CD4 T cell differentiation status in glaucoma patients

Findings from these PBMC transfer experiments suggest that functional differences exist in the leukocyte populations between POAG patients and controls. To examine whether these are reflected in alterations in distribution of T cell subsets, differentiation status, and activation status between POAG patients and controls, we optimized a 29-color spectral flow cytometry panel to comprehensively examine the T cell compartment. This assay is capable of identifying more than 16 different subsets of T cells and has the capability to distinguish the differentiation and activation status of these subsets (43). We used this panel to investigate PBMCs from glaucoma patients with severe disease (n=27), POAG patients with mild/moderate disease (n=19) and healthy controls (n=38).

Our initial flow cytometry gating strategy for PBMCs was as previously described (43). Lymphocytes were first identified based on forward scatter (FSC-A) and side scatter (SSC-A). Doublets were excluded by gating on single cells (FSC-A vs. FSC-H). Live CD45⁺ leukocytes were then selected using viability dye exclusion, followed by identification of CD3⁺ T cells. CD3⁺ cells were further subdivided into CD4⁺ and CD8⁺ T cell populations (Supplementary Figure 1A). Within the T cell compartment, we did not observe any significant differences in the frequencies of total CD3^+^ T cells, CD4 T cells or CD8 T cells (Supplementary Figure 1B).

Conventional CD4^+^ T cells were identified by expression of CD127 and exclusion of the transcription factor FoxP3 (Figure 3A). CD4s were further categorized based on CD45RA and CCR7 expression into naïve, central memory, and effector memory populations. The overall frequencies of naïve (p = 0.3227), central memory (p = 0.3614), and effector memory (p = 0.3517) CD4⁺ T cells did not differ significantly among healthy controls, mild/moderate POAG, and severe POAG patients (Figure 3B) We next performed further stratification of effector memory (Q4) CD4⁺ T cells based on CD27 and CD28 expression to define intermediate effector (Q1), early effector (Q2), early-like effector (Q3), and terminal effector (Q4) subsets (Figure 3C). This revealed selective alterations in effector differentiation states. Intermediate effector cells (Q1) were significantly increased across clinical groups (p < 0.0001), and early-like effector cells (Q3) also differed significantly (p = 0.0067). In contrast, early effector (Q2; p = 0.8422) and terminal effector (Q4; p = 0.2502) subsets were not significantly different (Figure 3D). Together, these findings indicate that while the overall distribution of major CD4⁺ T cell memory compartments remain stable in glaucoma, specific effector differentiation states within the effector memory pool are selectively altered, suggesting shifts in CD4⁺ T cell functional maturation in disease.

**Figure 3:**
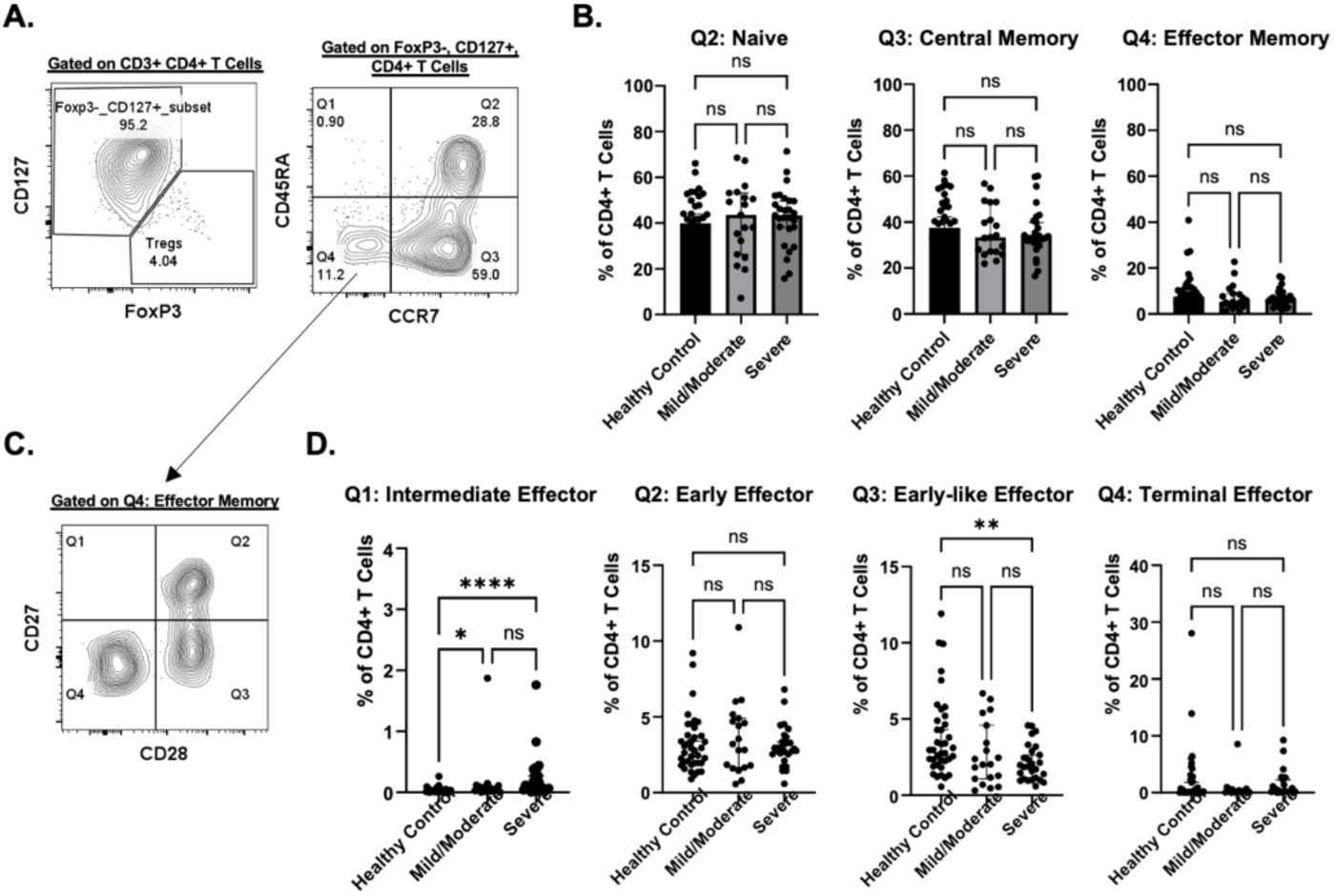
Altered CD4 T cell differentiation status in glaucoma patients. (A) Representative gating strategy. CD3+CD4 + T cells were gated, followed by exclusion of FoxP3 + CD127low regulatory T cells. Conventional CD4 + T cells were then subdivided based on CD45RA and CCR7 expression into naïve (Q2), central memory (Q3), and effector memory (Q4) populations. (B) Naïve (p=0.3227), central memory (p=0.3614), and effector memory (p=0.3517) CD4 + T cell frequencies in respective cohorts. Data are shown as % of CD4 + T cells. (C) Characterization of effector memory (Q4) CD4 + T cells based on CD27 and CD28 expression to define intermediate effector (Q1), early effector (Q2), early-like effector (Q3), and terminal effector (Q4) subsets. (D) Frequency of differentiated effector CD4 T cell subsets across clinical groups: Q1 p p<0.0001, Q2 p=0.8422, Q3 p=0.0067, and Q4 p=0.2502. Data are presented as % of CD4 + T cells. Statistical comparisons were performed using Kruskal-Wallis One-Way ANOVA test followed by Dunn’s multiple comparisons test as displayed on graphs: ns, not significant, *p<0.05, **p<0.01, ***p<0.001, ****p<0.0001.

### Disease–Associated Changes in Treg Subset Composition in POAG Patients

To determine if there are disease–associated changes in Treg subsets in POAG patients, we performed detailed phenotypic analysis of peripheral CD4⁺ Tregs. Tregs were subdivided into naïve (CD45RA^high^CCR4^low^), memory (CD45RA^low^CCR4^high^), and activated (CD39 ^high^HLA-DR^high^) populations (Figure 4A). Among the samples analyzed, the overall frequency of total Tregs among CD4⁺ T cells did not differ significantly across healthy controls, mild/moderate POAG, and severe POAG patients (p = 0.8121). However, subset analysis revealed significant disease-associated shifts. Naïve Tregs were significantly increased in glaucoma groups compared to controls (p < 0.0001). Conversely, memory Tregs (p < 0.0001) and activated Tregs (p < 0.0001), were reduced in POAG patients regardless of disease severity (Figure 4B).

**Figure 4:**
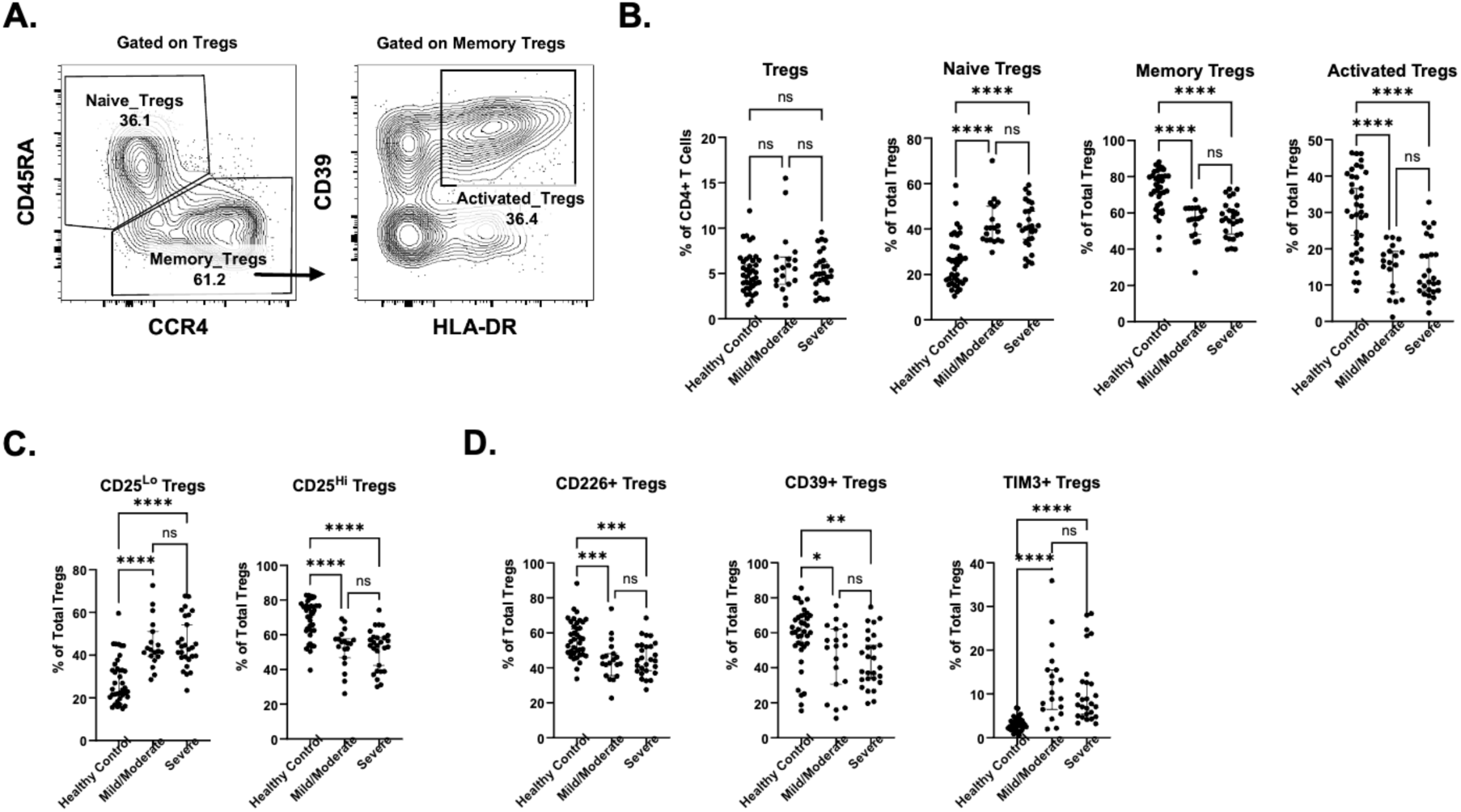
Disease–Associated Changes in Treg Subset Composition in POAG Patients. (A) Representative gating strategy for regulatory T cell (Treg) subset identification. Tregs were subdivided into naïve (CD45RAHighCCR4Low), memory (CD45RALowCCR4High), and activated (CD39HighHLA-DRHigh) populations. (B) Frequency of total Tregs (p=0.8121), naïve Tregs, memory Tregs, and activated Tregs, in healthy controls, mild/moderate POAG, and severe POAG patients. (C–D) Frequency of phenotypically distinct Treg subsets including CD25low, CD25High, CD226+, CD39+, and TIM-3+ across clinical groups. Data are shown as % of total Tregs. Statistical comparisons were performed using Kruskal-Wallis One-Way ANOVA test followed by Dunn’s multiple comparisons test as displayed on graphs: ns, not significant, *p<0.05, **p<0.01, ***p<0.001, ****p<0.0001.

Further Treg characterization demonstrated significant differences in functionally distinct Treg subsets (Figure 4C–D). We observed an expanded proportion of CD25^low^ (p < 0.0001) Tregs and correspondingly a reduced population of CD25^high^ (p < 0.0001) in POAG groups compared to controls which may indicate a loss of regulation function. Additionally, expression of activation and exhaustion-associated markers were altered: CD226⁺ Tregs (p < 0.0001), CD39⁺ Tregs (p = 0.0023), and TIM-3⁺ Tregs (p < 0.0001). These findings indicate that although total Treg frequency remains stable in POAG, the composition and activation state of Treg subsets are significantly remodeled in POAG, suggesting functional reprogramming of the regulatory compartment.

### Unsupervised Clustering Reveals Distinct Treg Subpopulations Associated with Clinical Status

While these initial analyses indicated that POAG patients exhibit abnormalities in the Treg compartment, they did not account for the functional differences observed between samples from patients with advanced versus moderate glaucoma following adoptive transfer into NSG mice. To better capture the phenotypic heterogeneity within the Treg compartment beyond conventional biaxial gating, we applied t-distributed stochastic neighbor embedding (t-SNE) visualization and unsupervised Phenograph clustering to identify disease-associated subpopulations in an unbiased manner. To prevent over-representation of any one sample in the downstream analysis, 1,000 Tregs from each donor were concatenated into a single WSP file. tSNE algorithm parameters utilized T cell markers including KLRG1, HLA-DR, TIM-3, CXCR5, CD45RA, CD49d, CCR4, CD73, CD25, CCR7, CX3CR1, CD28, PD1, CD226, CXCR3, CD95, CD39, CD27, and CD38 with iteration=3,000, perplexity=50 and learning rate=8,925 (44). The resulting 2-dimentional plot demonstrated heterogeneity across the Treg population (Supplementary Figure 2A-B).

Unsupervised clustering using the Phenograph algorithm identified 16 distinct Treg subpopulations which were then compared across clinical groups (severe, mild/moderate, and healthy controls) (Supplementary Figure 2C). There was clear spatial segregation of Treg clusters with distinct distribution patterns across clinical groups (Figure 5A). Six of these clusters showed significant disease-associated shifts (Figure 5B). Three populations (Populations 1, 3 and 10) were significantly expanded in both mild/moderate and severe POAG cases compared to controls. Population 9 was also significantly increased in severe cases, but there was no significant difference between mild/moderate cases and controls (Figure 5B).

**Figure 5:**
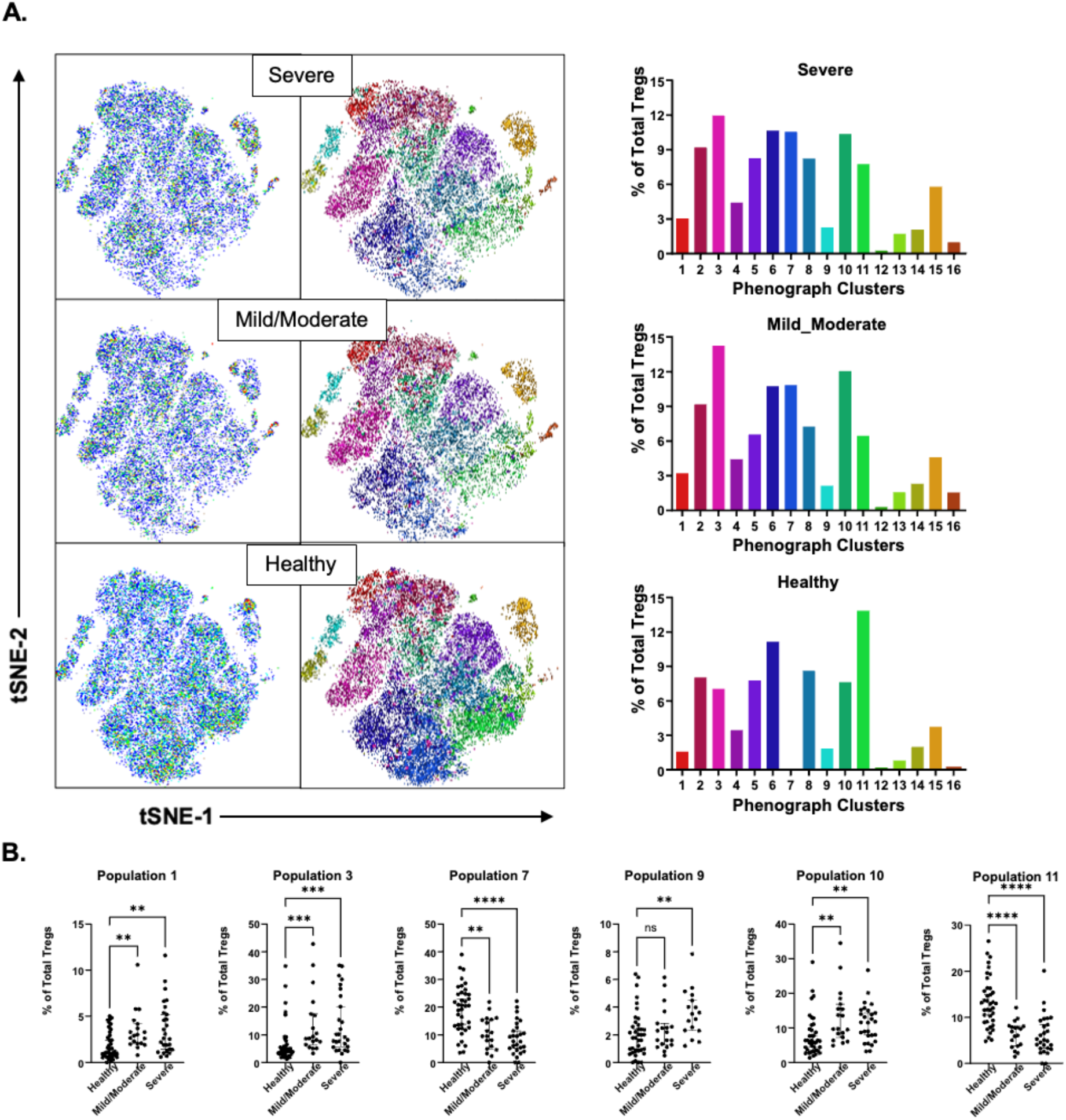
Unsupervised Clustering Reveals Distinct Treg Subpopulations Associated with Clinical Status. (A) tSNE map colored by Phenograph-defined clusters demonstrating spatial separation of Treg subpopulations for Healthy controls, Mild/Moderate and Severe POAG populations. Bar graph shows the frequency (% of total Tregs) of each Phenograph cluster across samples, highlighting differential enrichment of specific clusters between clinical groups. (C) Frequency of Phenograph-defined Treg subsets across clinical groups. Statistical comparisons were performed using Kruskal-Wallis One-Way ANOVA test: Population 1, Population 3, Population 7, Population 9, Population 10, and Population 11. Dunn’s multiple comparisons post-tests as displayed in graphs: ns, not significant, **p<0.01, ***p<0.001, ****p<0.0001.

Despite representing distinct differentiation states, all four of these populations share a central/lymphoid-homing phenotype characterized by CCR7 expression and lack of CX3CR1 which suggests they have a reduced capacity for inflammatory-site homing and tissue infiltration. Populations 1, 3 and 10 also exhibited reduced expression of the ectonucleotidases CD39 and CD73 which suggest partial or reduced adenosinergic suppressive capacity. Population 1 is a naïve/resting central Treg subset (CD45RA⁺CCR7⁺CD25⁺) with minimal activation or exhaustion marker expression. Population 9 is also a naïve/resting Treg population but has high expression of CXCR5 which is consistent with a resting, follicular-oriented central Treg that recirculates within lymphoid tissues rather than trafficking to inflamed peripheral sites. Population 3 is an effector memory Treg subset with evidence of prior activation (CD95^high^, CD27⁺, CD28^high^) but exhibits limited CD39/CD73-mediated adenosinergic capacity, consistent with recirculating central effector Tregs. It also has low expression of the marker CD38 which further implies a reduced suppressive capacity. Population 10 is Th1-polarized early effector central Treg (CD45RA⁺CCR7⁺HLA-DR⁺PD-1⁺CXCR3^hi^) yet retains a lymph-node homing (CCR7⁺) signature (Figure 5B, Figure 6 and Table 2). Populations 7 and 11 were significantly reduced in glaucoma patients (mild/moderate and severe) compared to healthy controls. They share a terminally differentiated, highly active effector Treg phenotype with strong tissue-homing and suppressive capacity. Population 7 is an antigen-experienced effector Treg subset (CD45RA⁻HLA-DR^high^CD95^high^) with strong suppressive potential (CD39^high^CD25^high^) and CCR4^high^CCR7^low^ expression which indicates tissue-homing capacity toward skin, mucosal and inflamed non-lymphoid sites. Population 11 has strong Th1-biased tissue-homing features (CXCR3^high^CCR4^high^CD49d^high^) and high suppressive function, which is consistent with a chronically stimulated, highly suppressive tissue-infiltrating effector phenotype (Figure 5B, Figure 6 and Table 2).

**Figure 6:**
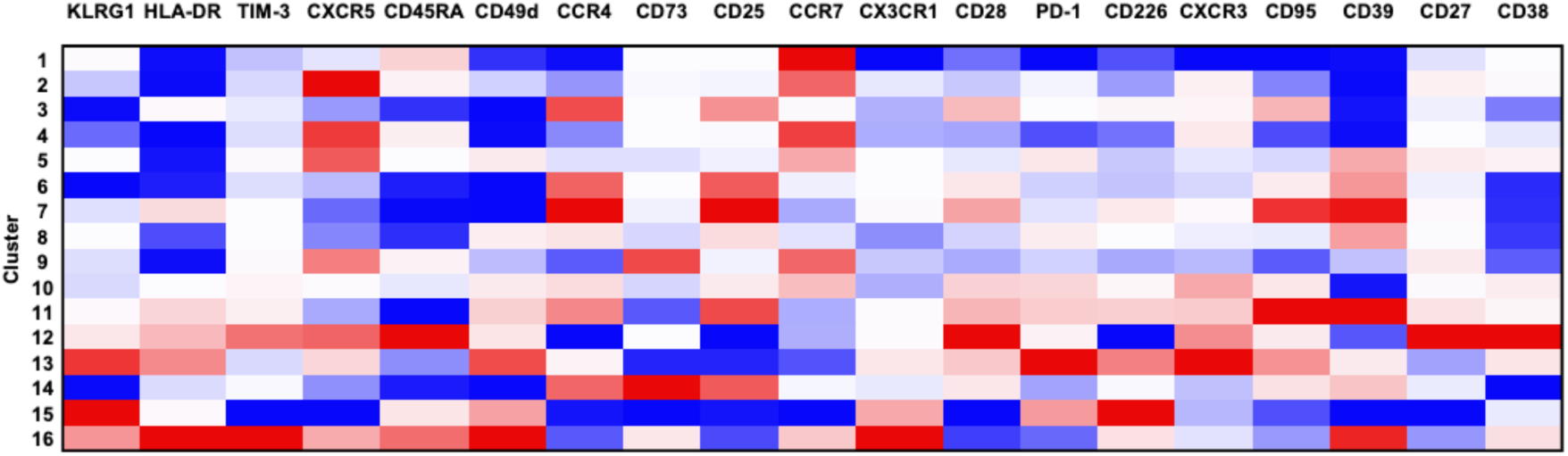
Heatmap of Treg Population Phenotypes Identified by Marker Expression. Heatmap showing relative expression of phenotypic, activation, homing, exhaustion, and suppressive markers across 16 Treg populations.

**Table 2:** Treg Population Phenotypes and Changes Across Glaucoma Disease Groups.

| Population # | Treg Subset | Trafficking Program | Activation/Exhaustion Profile | Regulatory Function | Frequency Trend across Disease Groups |
| --- | --- | --- | --- | --- | --- |
| 1 | Resting/naïve central Tregs | lymph node/central-memory homing | Low activation/Quiescent state | Low regulatory function | Significantly increased in glaucoma groups |
| 3 | Classic effector memory Tregs | lymph node/central-memory homing | Activated but not exhausted | Low regulatory function | Significantly increased in glaucoma groups |
| 7 | Highly activated, antigen-experienced effector Treg | Tissue homing capacity | Tregs are activated but not terminally differentiated or exhausted | Strong regulatory function | Significantly reduced in glaucoma groups |
| 9 | Resting/naïve central Tregs | Strong lymph node follicular/T-zone homing signature | Low activation/Quiescent state | Competent classical and adenosinergic suppressive capacity | Significantly increased in severe, but not mild/moderate glaucoma groups |
| 10 | Th1-polarized early effector central Treg | Strong Th1-trafficking signature with preserved lymph-node homing | Early activation, not exhausted | Low regulatory function | Significantly increased in glaucoma groups |
| 11 | Highly activated effector Tregs, with features of chronic activation | Strong Th1-trafficking signature with tissue infiltrating capacity | Highly activated but showing signs of exhaustion | Competent classical and partial adenosinergic suppressive capacity | Significantly reduced in glaucoma groups |

Together, these findings suggest that glaucoma is associated with an expansion of lymphoid-restricted, central Tregs with impaired suppressive capacity along with a reduction in highly suppressive, tissue-infiltrating effector Tregs. This indicates reduced peripheral regulatory control and altered Treg trafficking dynamics in disease.

### Expansion of CD25+FOXP3+ Tregs in Lag3^fl/fl.^Foxp3Cre^ERT2-GFP^ B6 mice after elevation of intraocular pressure reduces visual impairment

These findings indicate that reduced Treg activity may contribute to glaucoma progression in POAG patients. Augmentation of Treg activity has been shown to provide neuroprotective and anti-inflammatory effects in central nervous system (CNS) disorders, including multiple sclerosis and experimental autoimmune encephalomyelitis (45). To directly test whether modulation of regulatory T cells influences glaucomatous neurodegeneration, we employed an experimental mouse model of IOP elevation in which Tregs can be selectively expanded. Lymphocyte activation gene-3 (Lag3) is an inhibitory co-receptor expressed on activated T cells and constrains T cell proliferation, expansion, and viability (46, 47). Conversely, genetic or pharmacological inhibition of Lag3 in Tregs has been shown to ameliorate autoimmune pathology in murine models of type 1 diabetes (48). Therefore, we used Lag3^fl/fl.^Foxp3Cre^ERT2-GFP^ B6 mice to transiently expand FOXP3^+^ Tregs (Supplementary Figures 3 and 4).

As Tregs have a limited life span and are continuously generated from FoxP3^-^ precursor cells, we expected that increased proliferation of Treg cells following tamoxifen induction would eventually decline as *lag3^-/-^*Tregs are replaced. We found that one week post intraperitoneal injection of tamoxifen, the proportion of CD25^+^FOXP3^+^ cells in the spleen and peripheral lymph nodes of mice increased reaching a peak in the second week (p=0.0046 and 0.046, respectively). By the third week post-injection, the proportion of Treg cells declined and resembled that of uninduced mice. Therefore, we treated the mice with tamoxifen every 4 weeks to maintain the increased proportion of CD25^+^FOXP3^+^ cells in the mice (Supplementary Figures 3 and 4).

Elevated IOP was induced in mice by injections of the adenoviral vector Ad5RSVmyocillinY437H (Ad5myoc) into the anterior chamber of the mouse eye, as previously described (16, 49) Lag3^fl/fl.^Foxp3Cre^ERT2-GFP^ B6 mice were divided into four groups (n=9 mice/group): naïve (no treatment), tamoxifen-control, High IOP and High IOP + Treg Boost. 4 weeks after induction of IOP, mice in the tamoxifen-control and IOP + Treg Boost groups received 75mg tamoxifen/kg body weight by intraperitoneal injection once every 24 hours for a total of 5 consecutive days^46^ and then monthly to maintain CD25^+^FOXP3^+^ expansion for the remainder of the experiment.

Within one week of viral vector injection, IOP increased approximately 7-12mmHg in the High IOP and IOP + Treg Boost groups and then remained stable until the end of the experiment (Supplementary Figure 5). Naïve and tamoxifen-control mice maintained stable baseline IOP levels throughout the study period. These results indicate that neither Treg expansion nor tamoxifen treatment alter the magnitude or duration of IOP elevation (Supplementary Figure 5).

To assess the visual function of these mice, we measured the optokinetic reflex (OKR) at baseline (prior to IOP elevation) and repeated measurements monthly (Figure 7A). Any mice with pre-existing visual deficits were excluded. Overall, Lag3^fl/fl^.Foxp3Cre^ERT2-GFP^ mice had an average visual acuity of 0.392±0.024 cycle/degrees (c/d), and this value remained essentially unchanged in the naïve, tamoxifen-control and IOP + Treg Boost groups over the course of the experiment. In contrast, the OKR of mice in the High IOP group decreased rapidly during the first month following IOP induction and then slowly decreased further to 0.322±0.026 c/d (p=0.001). The High IOP + Treg Boost group experienced a similar extent of OKR decline as the High IOP group in the first month, but after receiving tamoxifen induction, the OKR recovered to an average of 0.367±0.039 c/d. At the 3-month time point, the High IOP + Treg Boost group had significantly higher OKR scores than the High IOP group (p=0.0093) and similar responses as naïve mice (p=0.2181, Figure 7B). These data suggest that expansion of Tregs may rescue visual function in mice even under conditions of continued IOP elevation.

**Figure 7:**
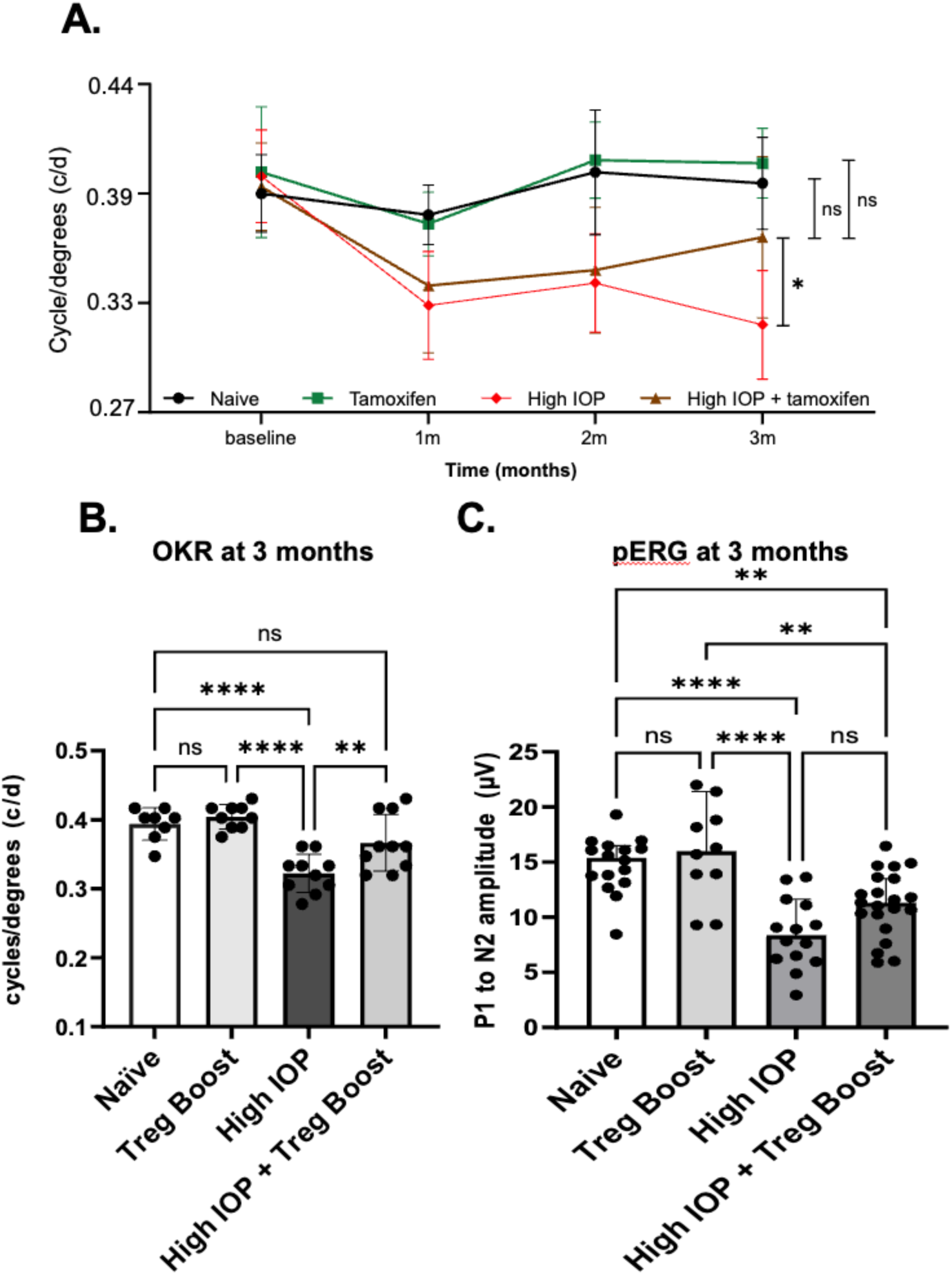
Treg Expansion Rescues Visual Function in Lag3^fl/fl^.Foxp3Cre^ERT2-GFP^ Mice following IOP Induction. (A) Optokinetic reflex (OKR; mean ± SEM, cycles) is shown at baseline and monthly time points after intraocular pressure (IOP) elevation. All groups exhibited comparable baseline values. Naïve (black circles) and Treg Boost (green squares) groups maintained stable visual function throughout the study. In contrast, the High IOP group (red diamonds) demonstrated a considerable decline one month after IOP elevation, with continued reduction over time. The High IOP + Treg Boost group (brown triangles) showed an initial decline like the High IOP group but exhibited significant recovery by the 3-month time point. (B) Quantification of OKR at 3 months. (C) For pattern electroretinography (pERG), the P1 to N2 amplitude is significantly diminished in High IOP group when compared to Naïve animals. The High IOP + Treg Boost group showed improved test outcomes, but statistical significance was not reached (p=0.078). One-way ANOVA, p<0.0001. Tukey’s multiple comparisons post-test: ns, not significant; *p<0.05, **p<0.01, ***p<0.001, ****p<0.0001.

To further evaluate RGC function in our mice, we recorded their retinal electrical activity via pattern electroretinography (pERG) during the 12th week of the experiment. The P1–N2 amplitude was comparable between naïve and tamoxifen-treated controls (14.81 µV vs. 15.88 µV; p = 0.83) and, as expected, was significantly reduced in untreated mice with elevated IOP (8.56 µV; p < 0.0001 vs. naïve). In High IOP + Treg Boost mice, pERG amplitudes improved (11.2 µV) relative to untreated mice, although this difference did not reach statistical significance (p = 0.078; Figure 7C).

### Treg Expansion Mitigates Axonal Damage and Preserves Peripheral Retinal Ganglion Cell Density in Experimental Glaucoma

Damage to the RGC axons in the optic nerve is a hallmark of glaucoma. In order to determine the number of damaged axons in each eye, one micron thick sections of optic nerves were prepared and treated with p-phenylenediamine (PPD) which preferentially stains disrupted myelin sheaths (Figure 8A). Quantitative analysis confirmed a significant increase in the number of damaged axons in the High IOP group relative to controls (p < 0.0001). Treg expansion significantly reduced axonal damage compared to untreated High IOP mice (p=0.0391), although damage levels were not completely ameliorated compared to controls (Figure 8B).

**Figure 8:**
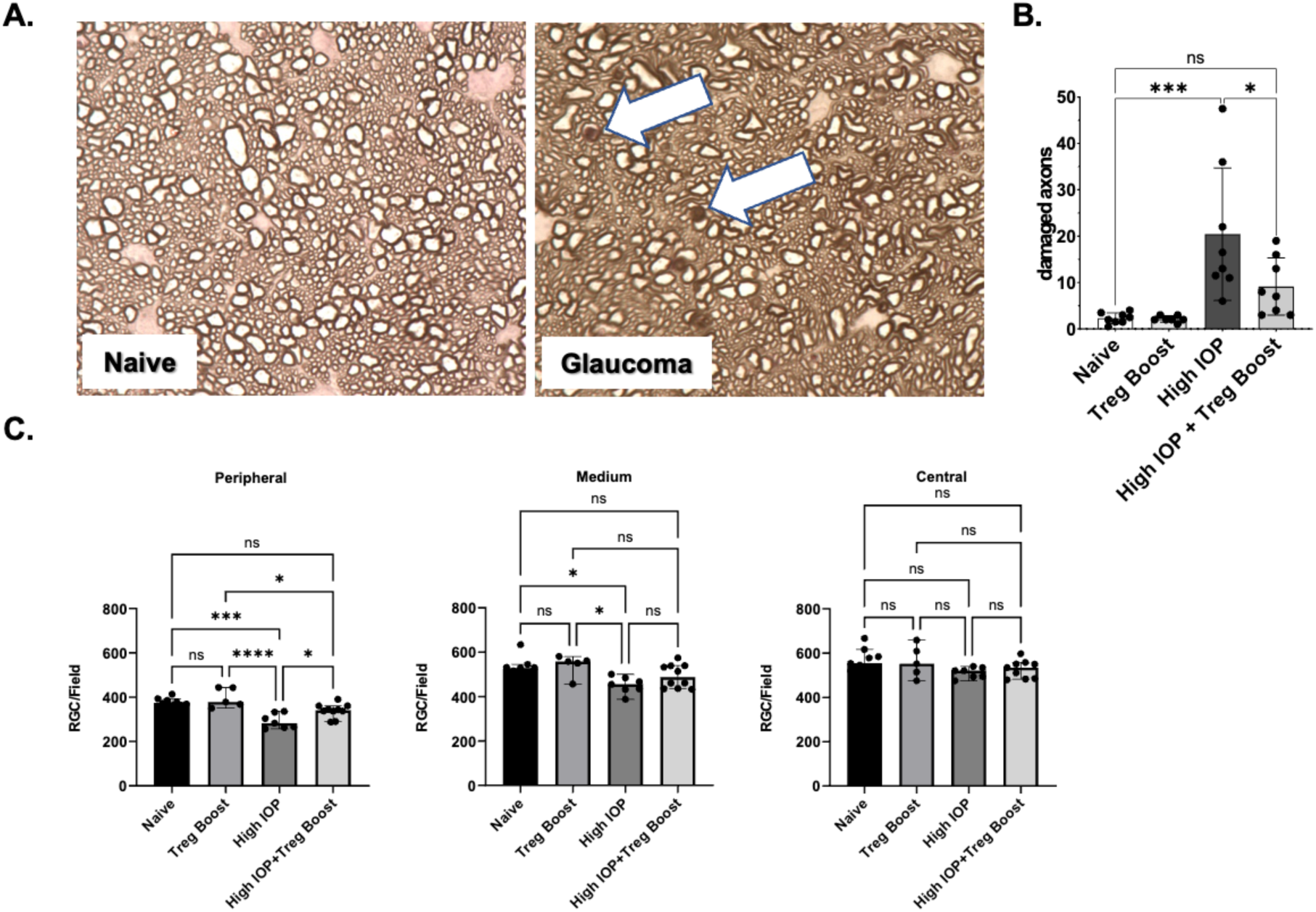
Treg Expansion Mitigates Axonal Damage and Preserves Peripheral Retinal Ganglion Cell Density in Lag3fl/fl.Foxp3CreERT2-GFP Mice following IOP Induction. (A) Representative optic nerve cross-sections from naïve and glaucomatous (High IOP) mice. Glaucomatous nerves show axonal degeneration that can be visualized by staining with p-phenylenediamine (PPD) (arrows) which is absent in naïve controls. (B) Quantification of damaged axons across experimental groups (mean ± SEM). The High IOP group exhibited significant increase in axonal damage compared to naïve and tamoxifen-only controls. Tamoxifen treatment significantly reduced axonal damage compared to untreated High IOP mice. (C) RGC density (cells/field; mean ± SEM) quantified in peripheral, mid-peripheral, and central retina. High IOP mice demonstrated significant RGC loss compared to naïve mice, particularly in the peripheral and mid-peripheral retina, while little damage was observed in the central region. All data were analyzed with one-way ANOVA with Tukey’s multiple comparisons post-test as shown on graphs: ns, not significant; *p<0.05, **p<0.01, ***p<0.001, ****p<0.0001.

While only very few axons are PPD positive in healthy optic nerves (Naïve: 2.3±1.2; Tamoxifen only: 2.1±0.7) these are significantly more numerous in animals exposed to elevated IOP (High IOP: 20.4±14.2 axons/section; p=0.0005 vs. naive). Concordant with the results of our functional analysis above, the optic nerves of mice in the High IOP + Treg Boost group displayed significantly fewer damaged axons than those of untreated High IOP mice (9.1±6.2 axons/section; p=0.039) indicating protection of the optic nerve against elevated IOP.

We next evaluated RGC density across retinal regions. High IOP mice exhibited significant RGC loss in the peripheral (p < 0.0001) and mid-peripheral retina (p = 0.0123), whereas RGC attrition in the central retina remained modest under these conditions (p = 0.1055; Figure 8C). Treg expansion preserved RGC density compared to untreated high IOP controls, with the most robust neuroprotection observed in the peripheral retina (p = 0.0296). No significant differences were observed in the central retina, likely due to the limited damage sustained in this region by the High IOP group. Collectively, these data demonstrate that Treg expansion mitigates axonal damage and preserves RGC density in experimental glaucoma, suggesting a neuroprotective role for regulatory T cells in IOP-induced degeneration.

## Discussion

We previously demonstrated in animal models that glaucoma elicits an adaptive immune response, primarily mediated by T cells, that is sufficient to drive RGC loss and may represent a pathogenic mechanism in the disease (16, 23, 24). Data from a number of clinical studies have indicated the presence of aberrant immune responses in POAG patients, suggesting that similar immune-mediated processes may also contribute to disease pathogenesis in humans (36–38).

In the present study, we provide direct functional evidence supporting this concept. Using a humanized NSG mouse model, we demonstrate that the transfer of PBMC from POAG patients induces significantly greater RGC damage in recipient mice compared with PBMCs obtained from control donors. These findings provide further confirmation that immune responses present in POAG patients are not merely correlative, but can actively mediate neurodegeneration, establishing a causal role for adaptive immunity in RGC destruction.

We initially focused on patients that presented to the ophthalmology clinic with POAG and optic disk hemorrhage, a finding strongly associated with future vision loss and active disease (50). In these patients and controls enrollment criteria were quite strict and individuals with any autoimmune or other neurodegenerative disorders were excluded. The resulting data clearly indicated an association of optic disk hemorrhage and the propensity of PBMC to cause RGC loss. Yet the majority of POAG patients do not have active disk hemorrhage and many glaucoma patients are afflicted with systemic conditions that could influence their immune function. We therefore expanded inclusion criteria in our second study to better reflect typical POAG patients. Data derived from this cohort also indicate that PBMC derived from POAG patients cause more RGC loss in recipient mice than those from non-glaucomatous controls. RGC loss was most pronounced in central portion of the recipient mice’s eyes. The reasons for this remain unclear, but it is worth noting that paracentral defects are also frequently observed in those patients developing glaucoma in the absence of elevated IOP (51). In these individuals, other disease mechanisms, which could include immune mediated damage, may be particularly important.

This study further demonstrates that the extent of RGC loss in NSG recipient mice is more pronounced in those animals that received PBMC from POAG patients with mild to moderate disease than those with advanced POAG. These findings align with recent data presented by Saini *et al*. demonstrating that Th1 cell responses to ɑ-crystallin are strongest in POAG patients with mild to moderate disease (52). Although the physiological basis and functional significance of this stage dependent activity remain unclear, the observations suggest that immune mediated mechanisms may be particularly relevant during the early phases of glaucomatous RGC degeneration. Notably, similar observations have also been made in other chronic neurodegenerative disorders, including Parkinson’s disease, in which inflammatory responses are highest in early disease and decline with progression (6, 53).

Taken together, these findings demonstrate that immune mediated processes capable of damaging RGCs are not confined to animal models of glaucoma but are also present in patients with POAG. Although the transferred PBMC cell population contains monocytes, dendritic cells, and lymphocytes, our previous studies in animal models strongly indicate a central role for T cells in mediating RGC damage in glaucoma (23, 24, 52). Yet mechanistic insight into the aberrant immune responses in glaucoma remains incomplete. To address this, we performed a comprehensive phenotypic analysis of the T cell compartment in POAG patients, including unconventional subsets and differentiation status.

While overall frequencies of major T cell populations were largely unchanged, multiple subsets exhibited a shift toward an early effector phenotype. T cell differentiation following activation is characterized by a defined loss of CD27 and CD28 expression and progressive acquisition of effector function. Such an enrichment of early effector T cells suggests persistent immune activation driving continued T cell differentiation, a hallmark of chronic inflammatory states (54–57). However, while the increased abundance of early effector T cells in POAG samples is suggestive of ongoing inflammation, our data does not distinguish whether this reflects immune activity specifically related to glaucoma or a generalized systemic inflammatory state.

While the changes in the differentiation status of effector T cell subsets in POAG patients are relatively modest, more pronounced differences were detected in the suppressive Treg compartment. Previous studies have reported both increases and decreases in the frequency of CD4 Tregs in POAG patients relative to healthy controls (36, 38). In the present study, we detected no differences in the overall frequency of CD4 Tregs; however, we observed a significant decrease in the proportion of activated Tregs in POAG patients when compared to healthy controls. Further, POAG patients exhibited an expansion of lymphoid-restricted, central Tregs with an impaired suppressive phenotype along with a reduction in highly suppressive, tissue-infiltrating effector Tregs. This indicates reduced peripheral regulatory control and altered Treg trafficking dynamics in disease.

The reduction in the proportion of activated Tregs was further accompanied by decreased expression of markers associated with Treg suppressive function. In particular, CD25 expression was significantly reduced in CD4+ Tregs from POAG patients. CD25, a high-affinity interleukin-2 (IL-2) receptor, is constitutively expressed by functional Tregs and enables efficient binding and internalization of environmental IL-2 (58, 59). Since IL-2 is required for T effector cells proliferation, sequestration of this cytokine is an important mechanism by which Tregs suppress immune responses. In addition, Il-2 signaling is also crucial for maintenance of FoxP3 transcription and therefore stability of Treg identity (29, 60). Consistent with these roles, multiple studies have demonstrated the CD25^low^FOXP3^+^ T cells are expanded in autoimmune diseases including diabetes, multiple sclerosis and systemic lupus erythematosus (61–64). Expansion of this population has also been described in age-associated immunosenescence (65, 66) and consequently loss of CD25 Tregs may be particularly relevant to age-related diseases such as glaucoma.

We also observed a reduced proportion of CD226+ cells within the CD4+ Treg population in POAG samples. Although multiple studies have reported that CD226 is associated with diminished suppressive capacity of CD4 Tregs (67–70), the role of CD226 in Treg biology is increasing recognized as context dependent. Notably, studies implicating CD226 signaling in reduced Treg suppression have utilized either CD226 knockout models or CD226 neutralizing antibodies (67–70). In contrast, CD226-specific ablation in CD4 Tregs has also shown to impair suppressive capacity, suggesting that CD226 is important for optimal CD4 Treg function (71, 72). In this context, the reduced frequencies of CD226⁺ and CD25+ CD4⁺ Tregs observed in POAG patients, together with the overall decrease in activated Tregs in this cohort, supports the notion of an unstable and functionally compromised Treg compartment in POAG, indicative of defective peripheral immune regulation.

These findings also suggest that expansion of Tregs may be able to mitigate glaucomatous neurodegeneration. Experimentally, Treg expansion can be achieved through exposure of T cell populations to rapamycin, an inhibitor of mTOR signaling that suppresses effector T cell proliferation while promoting FOXP3 expression and Treg survival (73). Although rapamycin has demonstrated neuroprotective effects in rodent models of glaucoma its broad effects on cellular metabolism and neuronal survival complicate discrimination between immunological and metabolic mechanisms (74, 75). To circumvent this limitation, we employed a genetic approach to selectively enhance Treg function by ablating the checkpoint inhibitor gene Lag3 specifically in Tregs, thereby promoting their expansion and suppressive activity upon activation (76). As reported previously, induction of elevated IOP in our animal model results in progressive visual dysfunction and structural degeneration (16, 23, 49, 77, 78). Notably, expansion of Tregs in mice with elevated IOP resulted in partial but significant preservation of visual function and reduced optic nerve damage compared with mice with intact *Lag3*. These results underscore that glaucoma is a multifactorial disease in which elevated IOP itself contributes directly to RGC damage, at least in mice, and are consistent with prior observations that elevated IOP induces less severe pathology in *Rag1*-deficient mice lacking adaptive immune cells (23). Together, our findings demonstrate that enhancing Treg function can ameliorate neurodegeneration in glaucoma, support a modulatory role for regulatory immune mechanisms in glaucomatous damage, and suggest that therapeutic strategies aimed at augmenting Treg activity may provide neuroprotective benefits to glaucoma patients.

One limitation of this study is that our analyses were performed using immune cells isolated from peripheral blood, which may not fully reflect immune processes occurring at sites of local inflammation. Although we have previously demonstrated T cell infiltration of the retina in POAG,(79) the phenotype and differentiation state of these tissue-resident cells remain unknown (24). As such, it is conceivable that the increased frequency of naïve and functionally impaired Tregs in the blood reflects preferential recruitment of activated or suppressive subsets to sites of inflammation. A second limitation is that immune cell populations were assessed as relative frequencies rather than absolute cell counts. While this approach is informative for identifying shifts in subset composition within an immune compartment, it does not allow determination of whether observed changes arise from true alterations in cell abundance or instead reflect proportional changes driven by expansion or contraction of other populations. The present work does not address the mechanisms underlying the emergence of autoreactive T cells targeting RGC. Autoreactive T cells are part of the normal peripheral repertoire and their frequency increases with age (80, 81). Under homeostatic conditions, their pathogenic potential is restrained by Tregs, but instability or dysfunction within that compartment may permit unchecked autoreactive T-cell activity, including responses that contribute to RGC degeneration.

A second limitation of this study is that the cohort was predominantly male and of Caucasian ancestry, reflecting the demographics of the patient population served at the two participating clinics. However, this distribution was similar in both POAG donors and controls, supporting that the observed association reflects a biologically meaningful role for diminished immune regulation in glaucoma that is detectable within this population. These findings provide a strong rationale for future studies aimed at validating and extending these observations in more diverse cohorts.

In conclusion, our presented data collectively provides evidence that dysregulation of an adaptive immune response in glaucoma is a central driver of RGC degeneration. The loss of Treg suppression as a potential failure of immune tolerance likely permits a glaucoma-specific immune component as another aspect to the multifactorial pathology. Our findings expand the current understanding of glaucoma beyond the known pathological mechanisms and impact of IOP elevation and puts glaucoma into the broader context of neurodegenerative disorders that are affected by immune mediated pathways. Our results clearly point to a dysregulation in Treg function and may offer a new avenue of treatment. Especially for glaucoma patients who continue to progress despite adequate IOP control, restoration of the Treg compartment presents a compelling and novel therapeutic approach.

## Methods

### Patient cohort

This study was approved by the University of Iowa Institutional Review Board (IRB #201711764, “Bioassay to Predict the Development and Progression of Glaucoma”) on February 7, 2018. Consented POAG patients and otherwise ocularly healthy controls were recruited from the Iowa City Veterans Affairs Medical Center (VAMC) and from the University of Iowa Department of Ophthalmology and Visual Sciences. Inclusion criteria involved a diagnosis of POAG, documented vision loss and changes to the optic nerve head. All participants were free of other severe ocular disorders or penetrating eye surgery (except cataracts), autoimmune disease, diabetes, current immunomodulatory therapy, or neurodegenerative disease. The age range was 53-91 years old with a mean age of 73±8.6 years and no statistical differences between the cohorts were observed (Table 1). PBMCs were obtained by venipuncture, isolated using Lymphoprep™ (StemCell, # 18061) and either immediately transferred to NSG mice by intraperitoneal injection or cryopreserved until further use.

### Animals

All animal experiments have been approved by the University of Iowa and the Iowa City VA IACUC and were conducted in accordance with the ARVO Statement for the Use of Animals in Ophthalmic and Vision Research. NOD.Cg-Prkdc^scid^ Il2rg^tm1Wjl^/SzJ (NSG) mice were obtained from The Jackson Laboratory (strain #005557). Lag3^fl/fl^.Foxp3Cre^ERT2-GFP^ B6 mice were a kind gift from Dr. D. Vignali (University of Pittsburgh) (76). All mice were housed in the animal facility of the Iowa City VAMC on a 12 h light–dark cycle (6 a.m. to 6 p.m.) with unrestricted access to food and water. All animals were euthanized by CO_2_ inhalation followed by cervical dislocation.

### Adoptive Transfer and Microbead Occlusion Model of Glaucoma

3.5 - 5 x 10^6^ PBMCs were transferred into male and female NSG mice by intraperitoneal injection. Each NSG mouse received PBMCs from a single patient and each patient sample was only injected into one mouse. Two weeks after PBMC transfer, IOP was elevated through the injection of 15 µm polystyrene microbeads into the anterior chamber. Prior to and after bead injection, IOP was recorded in 2.5% isoflurane-anesthetized mice using a rebound tonometer (Tonolab, Colonial Medical Supply, Windham, NH, USA). Only animals that showed IOP elevation between 17 and 20 mmHg that was resolved within 2 weeks were included in the study. Mice that exhibited signs of graft vs. host disease were excluded from the study.

Thirty days after IOP elevation, mice were euthanized, and the retinas were dissected. After fixation and dehydration, the retinas were immunostained with rabbit anti γ-synuclein antibodies (1:100 dilution, Abcam ab55424, Cambridge, MA, USA) following incubation in a donkey anti-rabbit Alexa Fluor 488 secondary antibody solution for three hours at room temperature in the dark or stained with 2% Nissl/Cresyl Violet to identify RGCs in the retinal ganglion cell layer (82). From each retina, 12 photomicrographs were taken at 400X at predetermined locations. RGC density was determined by investigators blinded to the status of the patient or mouse. One eye from each animal was used for RGC density analysis.

### Transient Treg Expansion in a Mouse Model of Elevated Intraocular Pressure

Elevated IOP was induced in Lag3^fl/fl.^Foxp3Cre^ERT2-GFP^ B6 mice by injections of the adenoviral vector Ad5RSVmyocillinY437H (Ad5myoc, University of Iowa Viral Vector Core, Iowa City, IA, USA) as previously described (16, 23). Briefly, newborn (P2-P4) mice received a subcutaneous injection of the adenoviral vector (3 × 10^3^ PfU) to induce tolerance to the vector, preventing ocular inflammation. At the age of 8 weeks, 9 × 10^7^ pfu virus in 3 µL PBS was introduced into the anterior chamber of the eye by transcorneal injection. The IOP of mice was measured by using a Tonolab tonometer between 9 am and 1 pm by a researcher who was blind to the animals’ treatment condition weekly. Animals that did not have an increase in IOP above the baseline by at least 7 mm Hg for at least 12 weeks were excluded from the analysis.

### PBMC immunophenotyping

Cryopreserved PBMC were quickly thawed in a 37°C water bath until ice crystals were no longer observed. PBMC were then poured from the cryovial into 10mL of pre-heated complete RPMI and live cells were counted by trypan blue exclusion. For surface staining, we employed a version of our optimized 31-color panel.^39^ Briefly, PBMC were plated at 1x10^6^ cells/well in a 96-well round-bottom plate and stained with Live/Dead Blue (ThermoFisher, Catalog # L23105) in 1x PBS for 15 minutes at room temperature. Cells were subsequently stained with antibodies in FACS buffer (2% FBS, 0.2% sodium azide in PBS) supplemented with Brilliant Stain buffer (BD Biosciences, Catalog # 566349) and blocking serum (rat, hamster, and mouse) for 30 minutes at room temperature. The surface marker antibodies and clones used were: anti-CD3 (SK7), anti-CD4 (SK3), anti-CD8 (SK1), anti-CD45RA (5H9), anti-CD11a (HI111), anti-CD49d (L25), anti-CCR4 (1G1), anti-CD73 (AD2), anti-CD27 (O323), anti-CCR7 (G043H7), anti-CX3CR1 (2A9-1), anti-CD28 (CD28.2), anti-PD-1 (EH12.2H7), anti-TIM-3 (7D3), anti-CXCR5 (RF8B2), anti-CD38 (S17015F), anti-CD226 (11A8), anti CXCR3 (G025H7), anti-CD95 (DX2), anti-TCRγδ (B1), anti-CD39 (A1), anti-KLRG1 (SA231A2), anti-CD127 (A019D5), anti-CD25 (BC96), anti-CD45 (2D1), anti-CD56 (NCAM16.2), anti-HLA-DR (L243). Following surface staining, cells were washed with FACs buffer and then resuspended in Foxp3 Fixation/Permeabilization buffer (ThermoFisher, Catalog # 00-5521-00) for 45 minutes at 4°C. Following fixation/permeabilization, cells were washed with Foxp3 permeabilization buffer and stained with anti-Foxp3 (PCH101) in permeabilization buffer at room temperature for 30 minutes. Cells were then washed twice with Foxp3 permeabilization buffer and resuspended in PBS. Cells were stored at 4°C until analysis.

For intracellular cytokine staining, PBMC were plated as above and stimulated with a cell stimulation cocktail (ThermoFisher, Catalog #00-4970-03) containing PMA, ionomycin, and protein transport inhibitors in complete RPMI for 4-5 hours at 37°C. As a negative control, a group of cells were resuspended in complete RPMI supplemented with only protein transport inhibitors (no stimulation). Following incubation, cells were stained with viability dye and surface markers as above, but with the following antibodies: anti-CD45RA (5H9), anti-CD8 (SK1), anti-CD3 (SK7), anti-CCR7 (G043H7), anti-TCRγδ (B1), anti-CD25 (BC96), and anti-CD45 (2D1). Cells were then washed with FACs buffer and resuspended in BD Cytofix/Cytoperm™ fixation and permeabilization solution (BD Biosciences, #554714) for 20 minutes at 4°C. Cells were then washed with and resuspended in BD permeabilization buffer (ThermoFisher, # 561651) supplemented with the following antibodies for 30 minutes at room temperature: anti-TNF (Mab11), anti-IL-10 (JES3-9D7), anti-IL-21 (3A3-N2), anti-IL-17A (BL168), anti-IFNγ (4S.B3), anti-IL-13 (JES10-5A2), anti-Granzyme B (QA16A02), anti-CD137 (4B4-1), anti-CD154 (TRAP1), and anti-CD4 (SK3). Cells were then washed with BD permeabilization buffer and then subsequently washed with and resuspended in PBS. Cells were stored at 4°C until analysis. Cells that were single stained with each of the antibodies stated above were used as compensation controls.

Data were collected on a Cytek® Aurora equipped with five lasers (355, 405, 488, 561, and 640nm) and 64 detectors and analyzed with SpectroFlo (Cytek Biosciences) and FlowJo software (FlowJo Version 10, Becton Dickinson, Ashland, OR USA).

For flow cytometry experiments involving LAG3 mice, splenocytes and lymphocytes were isolated and prepared as previously described. Staining cells with the following antibodies purchased from BioLegend (San Diego, CA): Percp-anti-mouse CD8 (Cat no:100732), Alexa Fluor 700-anti-mouse CD4 (Cat no:100430), PE/Cyanine7-anti-mouse CD3 (Cat no:100220), PE-anti-mouse FOXP3 (Cat no: 126404). APC-anti-mouse CD25 (Cat no:18-0251-82) purchased from ThermoFisher Scientific (Waltham, MA). Analysis of the flow cytometry data was done using FlowJo software.

### tSNE, Phenograph and ClusterExplorer

Using FlowJo, up to 1,000 Tregs from each donor were concatenated into a single WSP file. tSNE algorithm parameters utilized T cell markers including KLRG1, HLA-DR, TIM-3, CXCR5, CD45RA, CD49d, CCR4, CD73, CD25, CCR7, CX3CR1, CD28, PD1, CD226, CXCR3, CD95, CD39, CD27, and CD38 with iteration=3,000, perplexity=50 and learning rate=8,925.^40^ Phenograph was used to delineate cell clusters in an unbiased manner by unsupervised nearest-neighbors grouping.^40^ Populations that had less than 200 cells total across all samples were eliminated from further analyses. Another FlowJo plugin, Cluster Explorer, was used to investigate populations found through clustering using interactive plots displaying population frequencies, marker profiles and heatmaps (FlowJo). Following clustering, the FlowJo-generated parameter SampleID (which is retained during concatenation and encodes the original donor identity for each event) was used to deconvolute the merged dataset and reassign cell events to their respective donor samples while retaining the newly assigned tSNE and Phenograph parameters. This enabled calculation of per-sample cluster frequencies, rather than pooled event-level proportions.

### Tamoxifen-induced conditional gene-knockout

Tamoxifen was dissolved in corn oil at a concentration of 20 mg/ml. Mice received 75 mg tamoxifen/kg body weight by intraperitoneal injection once every 24 hours for a total of 5 consecutive days.^46^ Throughout the course of tamoxifen injections and for 72 hours thereafter mice were monitored daily for any adverse reactions to the treatment including dehydration, inability to reach food, hunched posture, reduced activity, and skin ulcers at the injection. Mice exhibiting distress or abnormal behavior were euthanized.

### Immunohistochemical Quantitation of Retinal Ganglion Cells

For γ-synuclein staining, thirty days after IOP elevation mice were euthanized, and the retinas were dissected. After fixation and dehydration, the retinas were immunostained with rabbit anti γ-synuclein antibodies (1:100 dilution, Abcam ab55424, Cambridge, MA, USA) following incubation in a donkey anti-rabbit Alexa Fluor 488 secondary antibody solution for three hours at room temperature in the dark^35^ or stained with 2% Nissl/Cresyl Violet to identify RGCs in the retinal ganglion cell layer.^80^ From each retina, 12 photomicrographs were taken at 400X at predetermined locations. RGC density was determined by investigators blinded to the status of the patient or mouse. One eye from each animal was used for RGC density analysis.

For RBPMS staining, mice were sacrificed, and immediately enucleated eyes were fixed in 4% paraformaldehyde for one hour. Blocked with 5% BSA/0.03% Triton-X100 for one hour at room temperature, retinas were incubated with rabbit-anti-mouse RBPMS antibodies (1:100 dilution, GeneTex, California, CA) at 4**°**C overnight on a rocker platform. After washes in PBS, binding was visualized following incubation in a donkey-anti-rabbit Alexa Fluor 488 secondary antibody solution for three hours at room temperature in the dark. Retinas were extensively washed in PBS and then mounted with Vectashield (Vector Laboratories, Burlingame, CA). RBPMS positive RGC were counted on twelve non-consecutive sections per eye.

### Pattern ERG (pERG) Recording

pERG recordings were performed as previously described (16). Mice were anesthetized by intraperitoneal injection of ketamine (30 mg/kg, Mylan, Canonsburg, PA, USA), xylazine (5 mg/kg, Akorn Inc., Lake Forest, IL, USA) and acepromazine (2.3 mg/kg, Rattlesnake Drugs, Scottsdale, Arizona, USA). After using 1% tropicamide to dilate the pupil, a drop of GenTeal gel (Alcon Laboratories, Fort Worth, TX, USA) was placed on the corneal surface to maintain corneal integrity. PERG was recorded using a Diagnosys Celeris System (Diagnosys LLC, Lowell, Massachusetts, USA) at 1 Hz (2 inversions per second) and 50 cd/m² alternating, reverse, black and white vertical stimulation response. 300 trajectories were recorded from each eye, the average waveform was calculated, and the amplitude (µV) from the peak of P1 to the valley of N2 is measured. All recordings were carried out while maintaining the animals’ body temperature between 37°C and 38°C using the system’s thermal pad.

### Optokinetic Reflex Measurements

Visual ability in B6 mice was measured using an OptoDrum (StriaTech, Tübingen Germany) as previously described (83). Mice are placed on a platform inside the apparatus, and a slowly rotating stripe pattern is generated on computer screens and presented to the animal. Reflexive tracking of the pattern is monitored by a camera and detected by the system’s software. Spatial frequency of the stimulus was stepped up or down with the staircase method to find the behavioral threshold. Lack of a tracking response indicates that the pattern is no longer perceived and defines the visual ability of the mouse. Trials in each session are repeated until the spatial frequency and direction can be determined unequivocally. In all cases the operator was blinded to the treatment status of the tested animal.

### Statistical analysis

Prior to statistical analysis, data were tested using the robust regression and outlier identification (ROUT) method using a Q equal to 1% (i.e. less than 1% of “statistically significant” outliers are false positives) (84). All statistically significant outliers were removed from analysis. Data were then assessed for normality using a D’Agostino and Pearson test. Data that were normally distributed were compared using a two-tailed t test. If an F test determined that the variances of the two groups were statistically different, a t test with Welch’s correction was used. *P* values for multi-group comparisons groups were calculated using One-Way ANOVA followed by Tukey’s post hoc tests. For data that were not normally distributed, data were compared using a Mann-Whitney test. Statistical multi-group comparisons were performed using Kruskal-Wallis One-Way ANOVA test followed by Dunn’s multiple comparisons test. All calculations were performed using GraphPad Prism 8.1 (GraphPad Software, San Diego, CA, USA) and *P* values < 0.05 were considered statistically significant.

## Data Availability

All data produced in the present study are available upon reasonable request to the authors

## Acknowledgement

This work was supported in part by Award I01 RX002860 and Center Award 1I50RX003002 from the United States (U.S.) Department of Veterans Affairs, Rehabilitation Research and Development Service. The contents do not represent the views of the U.S. Department of Veterans Affairs or the United States Government.

## Notes

This work was supported by NIH Award Number R01EY034534, and VA RRD Awards I01 RX002860 and 1I50RX003002.

### Competing Interest Statement

The authors have declared no competing interest.

### Author Declarations

IRB of University of Iowa gave ethical approval for this work

