## Supplemental figure 1 for "Impaired regulatory T-cell–mediated immune tolerance promotes neurodegeneration in glaucoma"

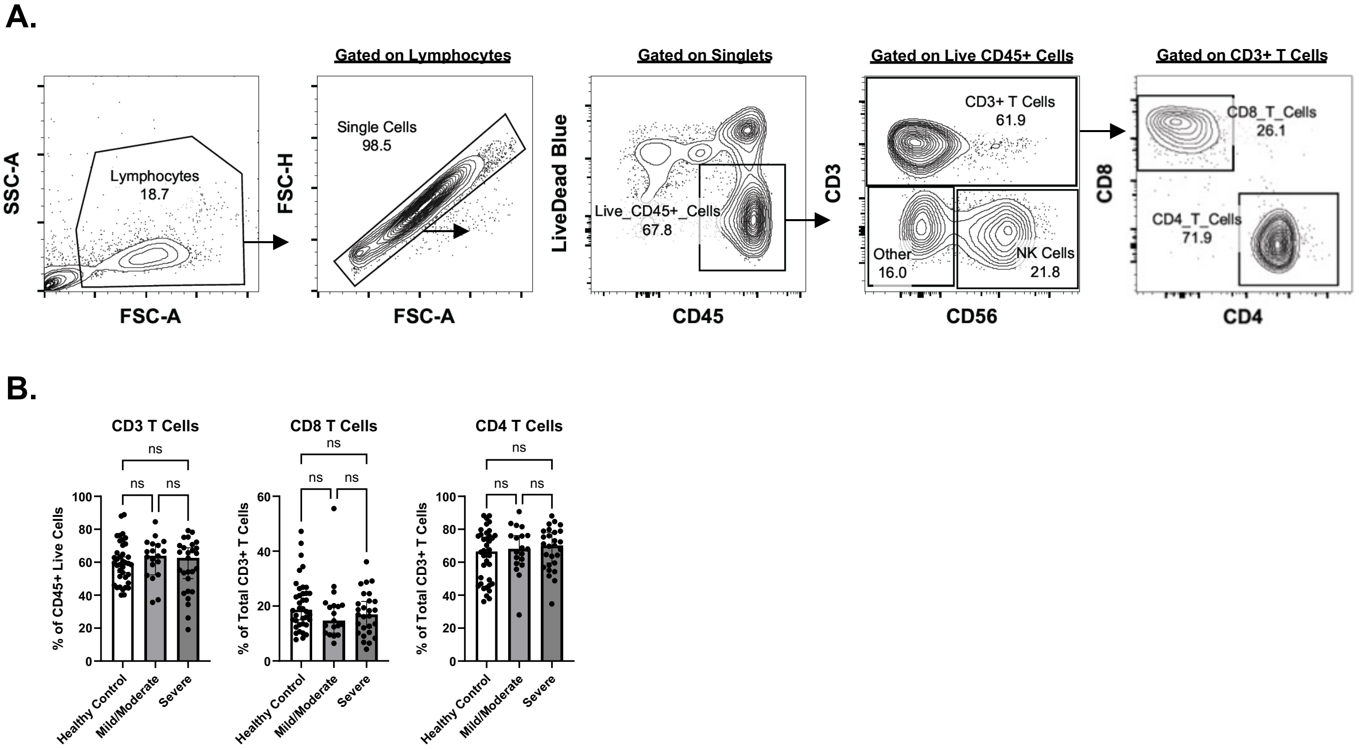


**Supplementary Figure 1: Flow cytometry gating strategy and peripheral T cell frequencies across disease severity groups.** (A) Representative flow cytometry gating strategy for peripheral blood mononuclear cells (PBMCs). Lymphocytes were first identified based on forward scatter (FSC-A) and side scatter (SSC-A). Doublets were excluded by gating on single cells (FSC-A vs. FSC-H). Live CD45$^+$ leukocytes were selected using viability dye exclusion, followed by identification of CD3$^+$ T cells. CD3$^+$ cells were further subdivided into CD4$^+$ and CD8$^+$ T cell populations. Representative percentages for each gate are shown within the plots. (B) Quantification of CD3$^+$ T cells (% of CD45$^+$ live cells), CD8$^+$ T cells (% of total CD3$^+$ T cells), and CD4$^+$ T cells (% of total CD3$^+$ T cells) in healthy controls, mild/moderate disease, and severe disease groups. Data are presented as mean ± SEM with individual data points shown. Statistical analysis via One-way ANOVA. No statistically significant differences were observed among groups (ns, not significant).
