## Supplemental figure 2 for "Impaired regulatory T-cell–mediated immune tolerance promotes neurodegeneration in glaucoma"

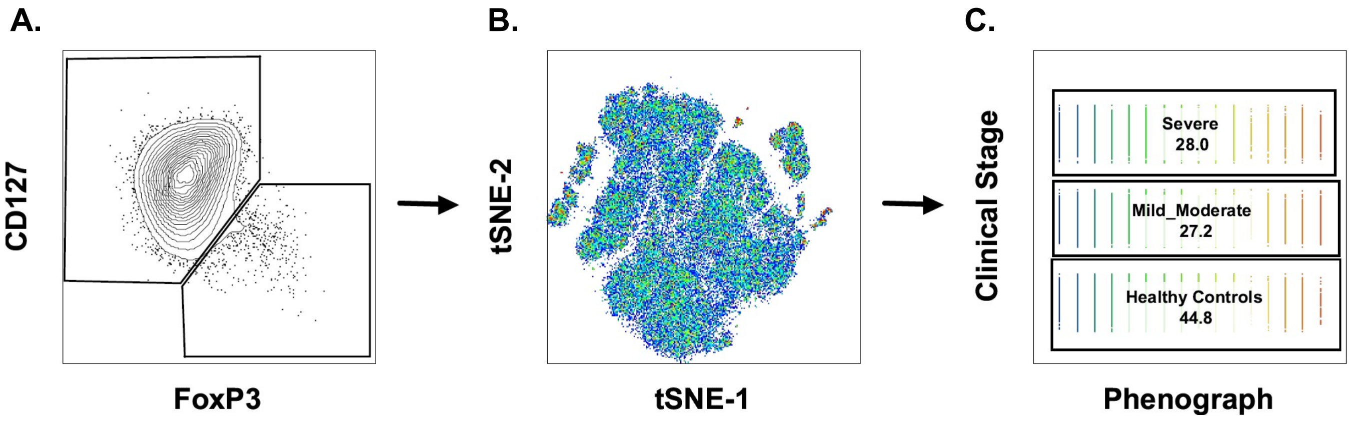


**Supplementary Figure 2: High-dimensional clustering to investigate clinical stage–associated changes in CD127⁻FoxP3⁺ regulatory T cells.** (A) Flow cytometric identification of regulatory T cells (Tregs) defined as $CD127^{low/-}FoxP3^{+}$ cells within the $CD4^{+}$ T cell compartment. Gated Tregs from all samples were concatenated for high-dimensional analysis. (B) t-distributed stochastic neighbor embedding (tSNE) projection of pooled Tregs from all subjects, demonstrating phenotypic heterogeneity across the Treg population. (C) Unsupervised clustering using the Phenograph algorithm identified distinct Treg sub-clusters which were then compared across clinical groups (Severe, Mild/Moderate, and Healthy Controls).
