## Supplemental figure 3 for "Impaired regulatory T-cell–mediated immune tolerance promotes neurodegeneration in glaucoma"

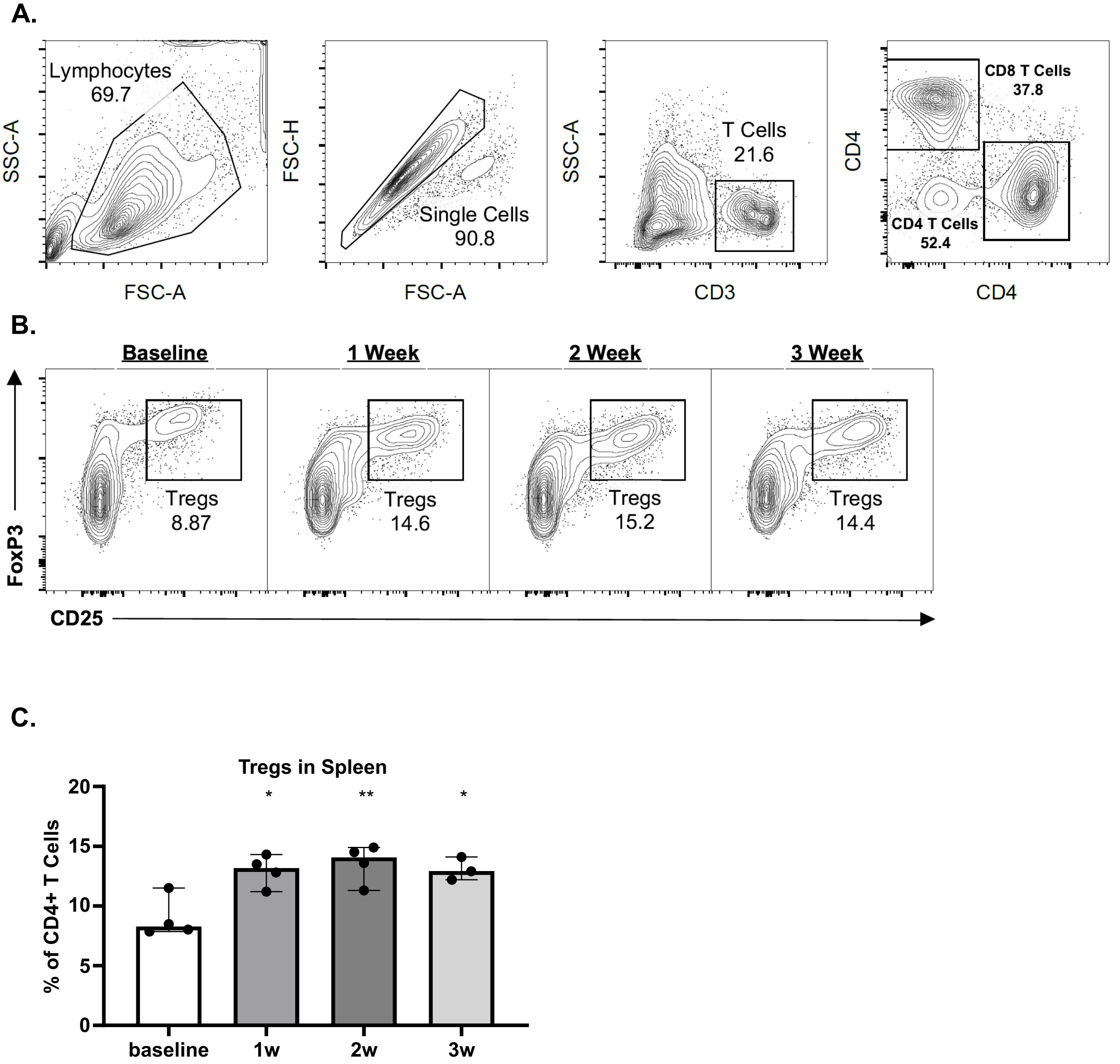


**Supplementary Figure 3. Expansion of Splenic Regulatory T (Treg) cells in Lag3$^{fl/fl}$.Foxp3Cre$^{ERT2\text{-}GFP}$ Mice Following Tamoxifen Treatment.** (A) Flow cytometry gating strategy for splenic T cell analysis. Lymphocytes were first identified based on FSC-A and SSC-A parameters, followed by exclusion of doublets using FSC-A versus FSC-H gating. CD3$^+$ T cells were gated and subsequently subdivided into CD4$^+$ and CD8$^+$ T cell populations. (B) Representative flow cytometry plots showing CD4$^+$CD25$^+$FoxP3$^+$ regulatory T cells (Tregs) at baseline and 1-, 2-, and 3-weeks post-treatment. The percentage of Tregs among CD4$^+$ T cells is indicated in each panel. (C) Quantification of splenic Tregs expressed as percentage of CD4$^+$ T cells (mean ± SEM). Treg frequencies increased at 1 and 2 weeks compared to baseline, with a sustained elevation at 3 weeks. Statistical significance One-Way ANOVA ($p=0.0036$) with Tukey's post-test multiple comparisons relative to baseline as indicated on graph (*$p < 0.05$, **$p < 0.01$).
