## Supplemental figure 4 for "Impaired regulatory T-cell–mediated immune tolerance promotes neurodegeneration in glaucoma"

A.

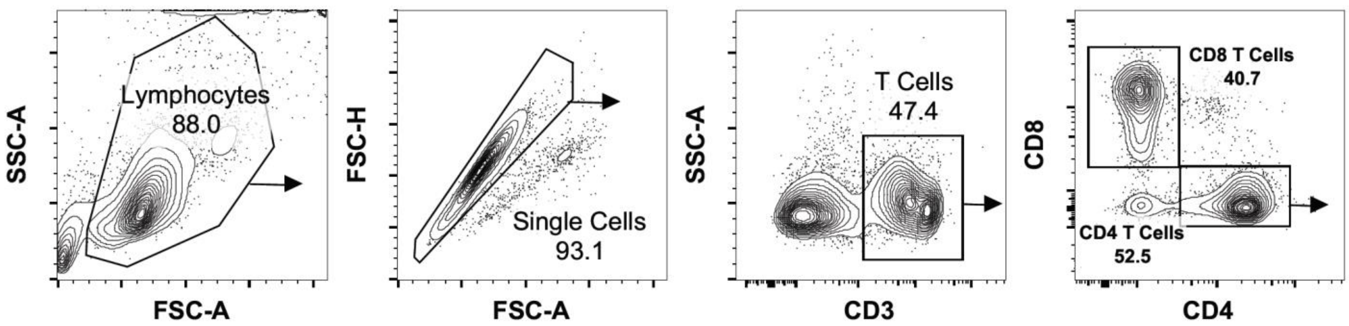


B.

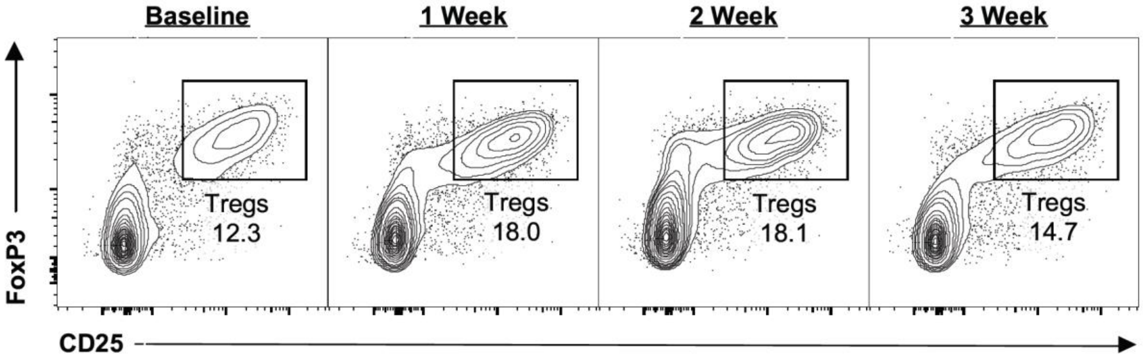


C.

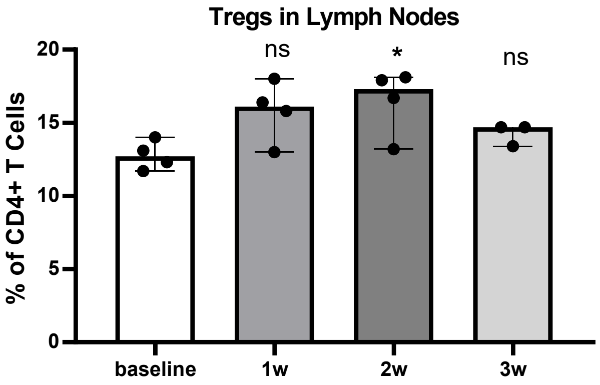


**Supplementary Figure 4. Expansion of Regulatory T (Treg) cells in Peripheral Lymph Nodes of Lag3$^{fl/fl}$.Foxp3Cre$^{ERT2\text{-}GFP}$ Mice Following Tamoxifen Treatment.** (A) Flow cytometry gating strategy for splenic T cell analysis. Lymphocytes were first identified based on FSC-A and SSC-A parameters, followed by exclusion of doublets using FSC-A versus FSC-H gating. CD3$^{+}$ T cells were gated and subsequently subdivided into CD4$^{+}$ and CD8$^{+}$ T cell populations. (B) Representative flow cytometry plots showing CD4$^{+}$CD25$^{+}$FoxP3$^{+}$ regulatory T cells (Tregs) at baseline and 1-, 2-, and 3-weeks post-treatment. The percentage of Tregs among CD4$^{+}$ T cells is indicated in each panel. (C) Quantification of peripheral lymph node Tregs expressed as percentage of CD4$^{+}$ T cells (mean ± SEM). Treg frequencies increased by 2 weeks compared to baseline, with contraction at 3 weeks. Statistical significance One-Way ANOVA ($p=0.0480$) with Tukey's post-test multiple comparisons relative to baseline as indicated on graph (ns, not significant; $^{*}p < 0.05$).
