## Supplemental figure 5 for "Impaired regulatory T-cell–mediated immune tolerance promotes neurodegeneration in glaucoma"

A.

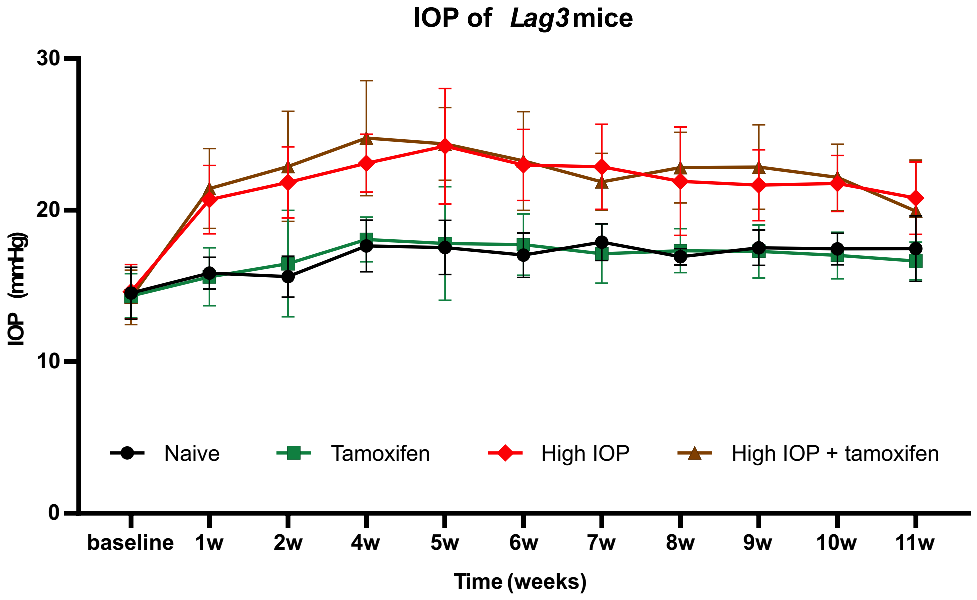


**Supplementary Figure 5: Intraocular Pressure of Lag3$^{fl/fl}$.Foxp3Cre$^{ERT2\text{-}GFP}$ Mice Over Time. (A)** Intraocular pressure (IOP) was measured longitudinally at baseline and weekly for 11 weeks in naïve, tamoxifen-only, High IOP, and Tamoxifen + High IOP groups. Naïve and Tamoxifen-only mice maintained stable IOP levels throughout the study period. In contrast, High IOP-treated mice exhibited a rapid increase in IOP beginning at 1 week, peaking around 4–5 weeks, and remaining elevated compared to controls. Data are presented as mean ± SEM (mmHg).
